# A high-resolution flux-matrix model describes the spread of diseases in a spatial network and the effect of mitigation strategies

**DOI:** 10.1101/2021.12.22.21268059

**Authors:** Guillaume Le Treut, Greg Huber, Mason Kamb, Kyle Kawagoe, Aaron McGeever, Jonathan Miller, Reuven Pnini, Boris Veytsman, David Yllanes

## Abstract

Propagation of an epidemic across a spatial network of communities is described by a variant of the SIR model accompanied by an intercommunity infectivity matrix. This matrix is estimated from fluxes between communities, obtained from cell-phone tracking data recorded in the USA between March 2020 and February 2021. We apply this model to the SARS-CoV-2 pandemic by fitting just one global parameter representing the frequency of interaction between individuals. We find that the predicted infections agree reasonably well with the reported cases. We clearly see the effect of “shelter-in-place” policies introduced at the onset of the pandemic. Interestingly, a model with uniform transmission rates produces similar results, suggesting that the epidemic transmission was deeply influenced by air travel. We then study the effect of alternative mitigation policies, in particular restricting long-range travel. We find that this policy is successful in decreasing the epidemic size and slowing down the spread, but less effective than the shelter-in-place policy. This policy can result in a pulled wave of infections. We express its velocity and characterize the shape of the traveling front as a function of the epidemiological parameters. Finally, we discuss a policy of selectively constraining travel based on an edge-betweenness criterion.

---

**W**Hen plague hit Florence in August 1630, the Florentine authorities made a number of high-stakes decisions which proved highly effective [1]. One of the reasons the Florence *Sanità* could organize this response was the ample time they had, forewarned as they were by the Milanese authorities in November 1629. Today’s public-health authorities work under much more compressed timescales, as evidenced by the SARS-CoV-2 pandemic. Long-distance travel radically changes the dynamics of spreading, which raises a number of questions about the spatial dynamics of transmission in modern times. Epidemic outbreaks in the last two decades have provided the scientific community with a wealth of material to study these questions, going beyond the classic Susceptible-Infected-Recovered (SIR) theory with perfect mixing [2–6]. Several studies have shown how the total epidemic size can be affected by factors such as inhomogeneity in transmission rates [7–14] or in the mode of transmission [15, 16]. Classically, motion of individuals was taken into account by introducing diffusion terms in the standard SIR equations, allowing the emergence of spatio-temporal patterns [17–20]. Recently, in the context of the SARS-CoV-2 pandemic, such approaches have been especially valuable in order to study the effect of containment policies such as lockdown and quarantine [21, 22]. These models are, however, limited in that they do not, in principle, take long-distance air travel into account. Several works have, therefore, considered disease spread in a network, typically constructed from air-traffic data [19, 23, 24], where edges can connect locations separated by large geographical distances. This approach can lead to very accurate predictions at the country scale [25] but predictions at finer scales remain challenging. Another study considering human mobility emphasized how spatial variation in public-health infrastructure reflected on epidemiological parameters can affect the dynamics of spread to different countries [26]. Data-based studies of epidemic spread and the impact of social distancing through the analysis of social-network structure have also been very informative [16, 27–29].

A recent paper by Chang *et al*. [30] obtained a model for the spatio-temporal spread of a disease at a high spatial resolution by using extensive mobile tracking information to identify physical interactions between individuals. Chang *et al*. showed that the actual spread of the SARS-CoV-2 epidemic can be well explained from the mobility data of individuals. The model relied on the simulation of interactions among individuals on a bipartite graph where nodes, representing locations at a very fine spatial resolution, are divided in two sets: Census Block Groups (residential areas) and Points Of Interest (non-residential), each of them having its own transmission rate. The model was fitted to reproduce known reported cases of COVID-19 in 10 metropolitan areas, and could then be used to make short-term predictions about the spreading or study the effect of different mitigation strategies.

Here we take an approach similar to that of Chang *et al*., using mobility data to calibrate a model for disease spread. However, we investigate this propagation at the scale of a large country, the USA, rather than metropolitan areas. Specifically, we introduce a spatial SIR epidemiological model in which effective transmission rates between *N* = 2^10^ communities are computed from mobility data of individuals belonging to these communities. We show that this model captures well the spread of the SARS-CoV-2 epidemic. Remarkably, we find that a simple model consisting of an interaction frequency dropping under the effect of lockdown, and of a single flux matrix encoding the travel of individuals, faithfully reproduces the reported cases of COVID-19 both globally and locally in each community. Strikingly, the SARS-CoV-2 epidemic spreads in a delocalized fashion, infecting distant communities very quickly. Moreover, an even simpler model with uniform transmission rates between the communities gives results very close to the model based on mobility data, emphasizing the prevalent role air travel played in the spread of the SARS-CoV-2 epidemic. We then study how interventions that change travel patterns can localize epidemics. In particular, we investigate the hypothetical effect of a policy preventing long-distance travel. In addition to “flattening” the curve, spreading through nearest-neighbor interactions creates traveling waves, which we characterize both analytically and numerically. These results allow us to discuss which interventions are more effective, limiting short-range contacts (a lockdown), or limiting long-range trips (a quarantine). We then propose an alternative mitigation strategy based on an edge-betweenness criterion.

## Model

We consider a metapopulation model of *N* communities numbered 1, 2,…, *N*. Denoting by *S*_*a*_, *I*_*a*_ and *R*_*a*_ the numbers of susceptible, infected and recovered individuals in community *a*, the standard SIR equations read:

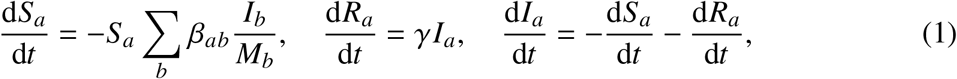

where *β*_*ab*_ is the transmission rate from infected individuals in community *b* to susceptible individuals in community *a* and *γ* is the recovery rate, assumed to be the same for all communities. Diagonal elements of the infectivity matrix [*β*_*ab*_] describe intracommunity infections, while off-diagonal elements describe inter-community infections (Figure 1**a-b**). We also introduce the local epidemic sizes *T*_*a*_ =*I*_*a*_ + *R*_*a*_. The total population in each community ***M***_*a*_ is constant through time,

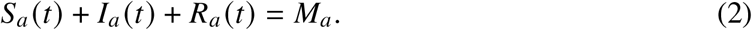

**Figure 1.**
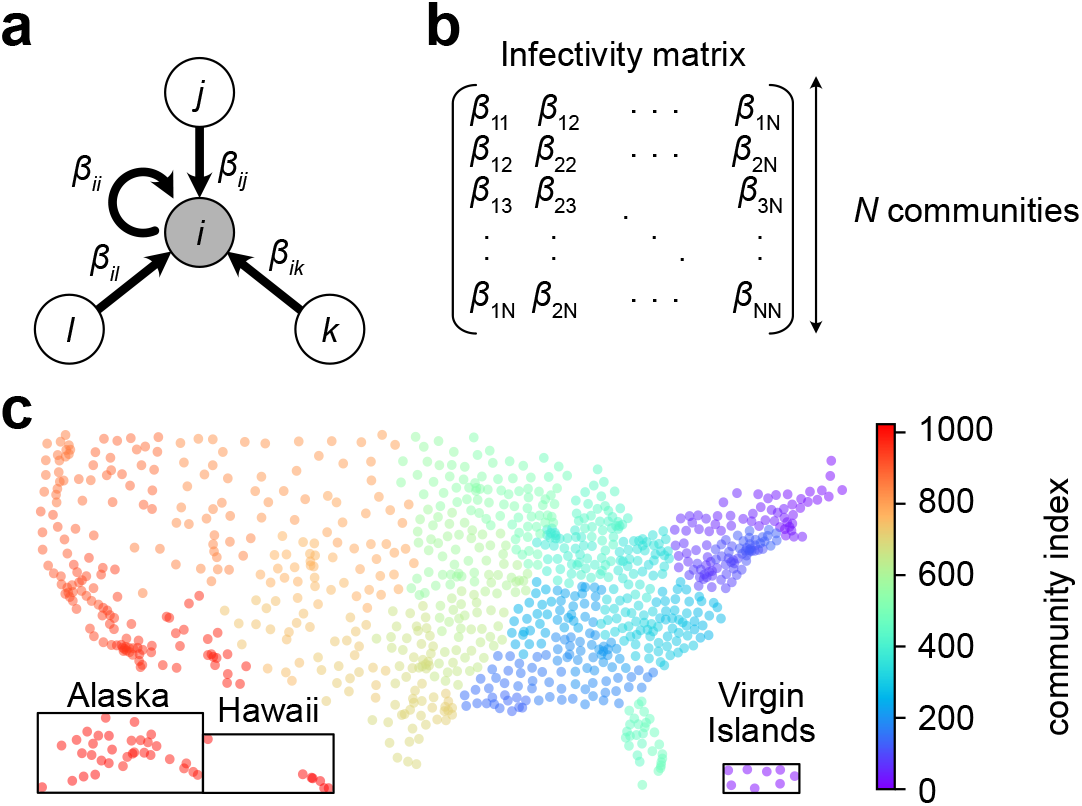
Model for the spreading of SARS-CoV-2 in a network of communities in the USA. **(a)**. One community *i* interacting with three other communities *j, k* and *l*, with the transmission rates *β*_*ij*_, *β*_*ik*_ and *β*_*il*_ respectively. **(b)** Infectivity matrix. **(c)** *N* = 2^10^ communities in the USA. Each community aggregates a number of Census Block Groups (CBGs).

The model in equation (1) has been extensively studied [31–35]. We show in the Supplementary Information (SI) that the dynamics can be reduced to an ODE of just one *N*-vector variable, and the endemic equilibrium can be obtained by solving a transcendental equation involving the infectivity matrix [*β*_*ab*_].

In order to estimate the infectivity matrix, we used mobility data compiled by SafeGraph [36], tracking the location of about 20 million USA cell phones between March 2020 and February 2021. The locations consist of more than 200 000 Census Block Groups (CBGs). Each cell phone is assigned for physical residence the location where it spent the most time, and daily visits to other locations are recorded. For computational purposes, we coarse-grained the physical locations into *N* = 2^10^ communities, which are shown in Figure 1**c**. Let *f*_*ab*_ be the number of individuals from community *a* visiting community *b* per unit of time. We will assume that

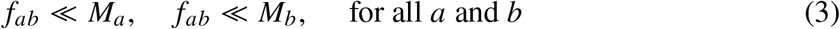

The variation in susceptible individuals in community *a* due to new infections during the time interval Δ*t* has the form:

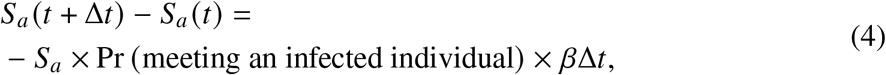

where *β* is the disease-specific transmission rate when a susceptible individual has contact with an infected individual. There are three kinds of infection to consider: (i) the infected person and the infector belong to the same community, (ii) an infected person visits a neighboring community and infects a resident of this community, and (iii) a susceptible person visits a neighboring community and gets infected by one of its residents. We will neglect the rarer “tourist to tourist” infections, when an infected person visits a neighboring community and infects there a visitor from yet another community. The term Pr meeting an infected individual can therefore be evaluated as a function of the pseudo-flux matrix [*f*_*ab*_] for each of the three aforementioned cases (SI). After summation of the three contributions, we obtain:

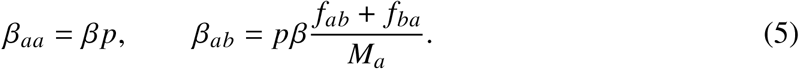

where *p* is the frequency with which an individual is having contact with an other individual of its community, and is assumed to be the same for all communities. Equation (5) determines the infectivity matrix [*β*_*ab*_] from equation (1) up to a proportionality constant, namely *βp*.

## Results

### The model reproduces the spatial dynamics

In order to assess the validity of the infectivity matrix based on mobility data, equation (5), we confronted the model’s predictions against COVID-19 case numbers reported in the USA by the Center for Systems Science and Engineering (CSSE) at Johns Hopkins University [37]. Specifically, we fitted the daily *βp*(*t*) values so as to minimize the sum of squared errors between values predicted by the model and values reported by the CSSE (Material and Methods). The fitted *βp* values show a steep decay during the month of March 2020, followed by a plateau lasting until February 2021 (Figure 2**b**). Although we fitted the model to the real COVID-19 confirmed cases using the global quantity Ω, local epidemic sizes _*a*_ predicted by the model agree very well to the empirical values.

**Figure 2.**
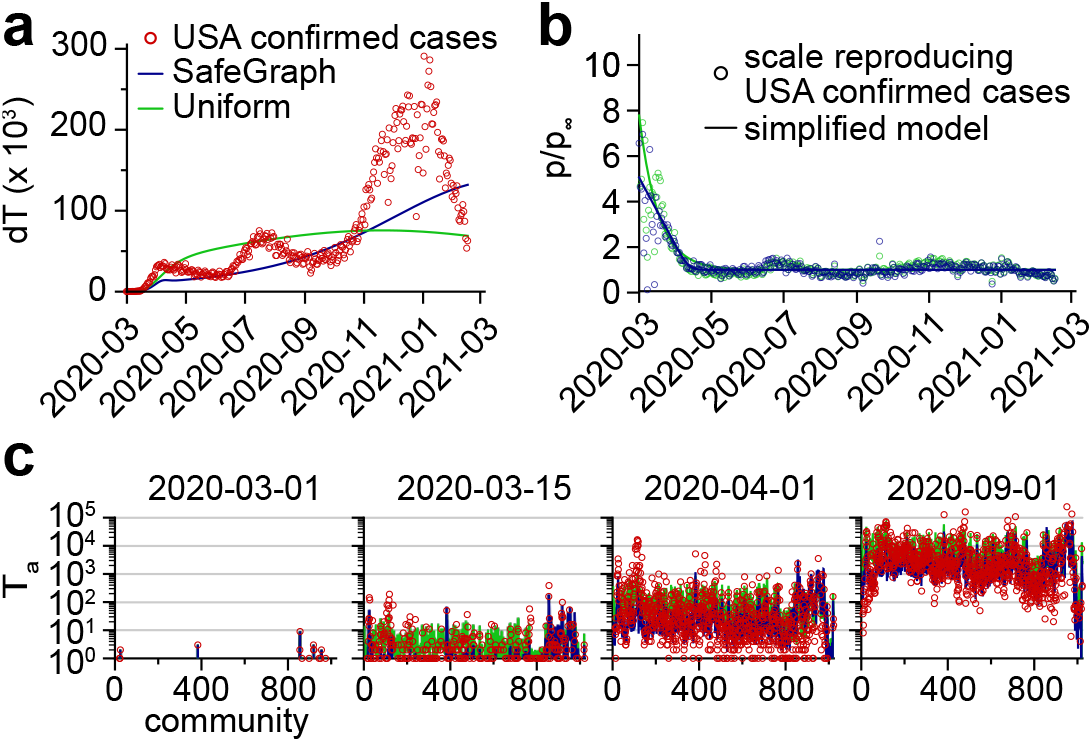
Model based on SafeGraph mobility data or a uniform infectivity matrix. **(a)** The model is fitted to COVID-19 confirmed cases in the USA. There is one fitting parameter per day. **(b)** Fitting parameters obtained. The shape suggests a simplified model with two limiting values before and after lockdown. **(c)** The simplified models obtained reproduce the spread of the SARS-CoV-2 epidemic in the community network. A direct comparison between the local epidemic sizes predicted by the model and the reported values can be found in Figure S1. See also Movies S1-2.

### A two-phase simplified model

The time variations of *βp*(*t*) shown in Figure 2**b** imply a drastic decrease in the interaction frequency among individuals across all communities. This result reflects the effect of the shelter-in-place policies that were implemented at the beginning of the SARS-CoV-2 pandemic in many USA states. We can use this observation to estimate the effect of these policies quantitatively: the interaction frequency among individuals, namely *p*, is about 5 times smaller in the plateau following shelter-in-place policies than it was at the onset of the SARS-CoV-2 pandemic. Following this observation, we defined a simplified model, in which the fit with reported cases was carried out while enforcing a softplus shape for *βp*(*t*) (Figure 2**b** and Material and Methods). This simplified model reproduced the average progression of the epidemic (Figure 2**a**), but it didn’t capture the three oscillations visible in the number of new cases. We conclude that these oscillations in the number of new cases were mostly driven by a similar oscillatory pattern in the interaction frequency as seen in Figure 2**b**. In Figure 2**c**, we show that the number of new COVID-19 cases in each community predicted by the simplified model follows closely the empirical values. This result suggests that the infectivity matrix constructed from mobility data (equation (5)) is a good approximation of the “true” infectivity matrix.

### Turning down long-range interactions

The previous results suggest that the decrease in new COVID-19 cases was mostly driven by a country-wide reduction in the interaction frequency among individuals. Here we investigate the hypothetical effect of an alternative policy, namely a travel restriction while keeping unchanged the interaction frequency among individuals. Specifically, we modified the infectivity matrix so that communities separated by a physical distance larger than a prescribed cutoff do not interact: *β*_*ij*_ = 0 if *d*_*ij*_ *> d*_*C*_ (Figure 3**a**). We seeded the infection in a community belonging to the state of Washington and simulated the spread of the epidemics using a fixed interaction frequency (*βp*(0) from the simplified model). As expected, we observed a reduction in the number of daily cases *dT* when the cutoff distance *d*_*C*_ decreased, illustrating the “curve-flattening” effect that was targeted by travel restriction policies (Figure 3**b**). In this idealized scenario with a single seed for the infection, the epidemic propagates as a traveling wave from the west coast to the east coast (Figure 3**c**). However, as can be seen by comparing Figure 2**a** to Figure 3**b**, travel restriction policies are not as efficient as lockdown policies to decrease the spread of an epidemic.

**Figure 3.**
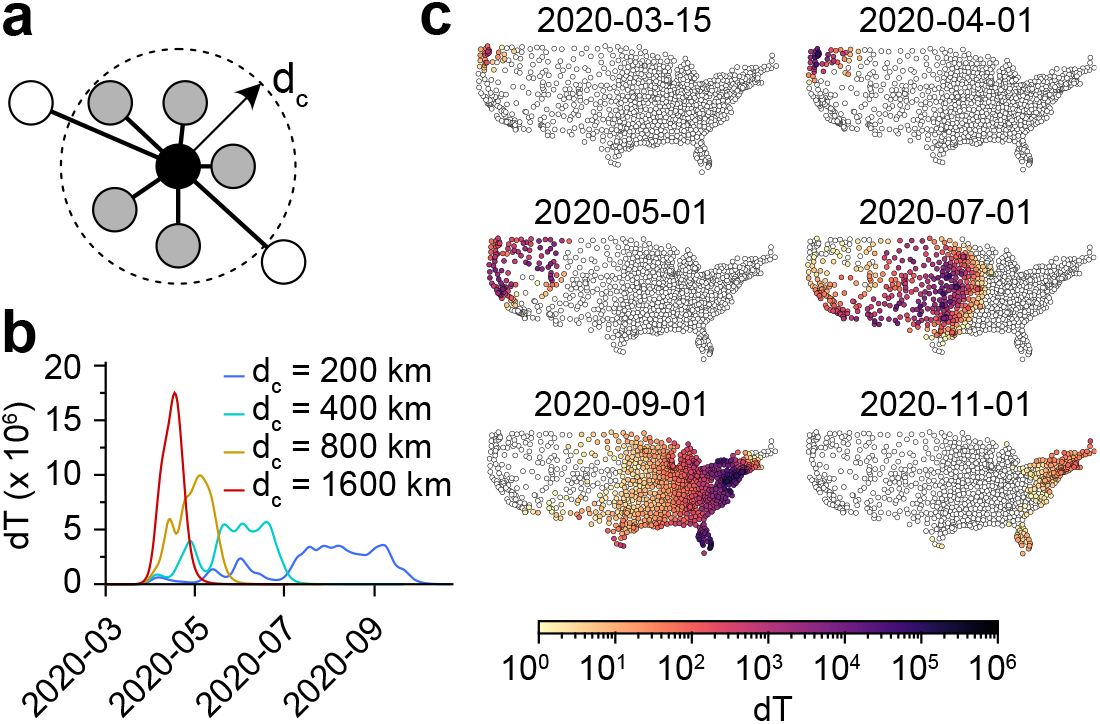
Limiting long-distance travels without local lockdown. **(a)** Only transmission rates for communities separated by a distance *d*_*ij*_ < *d*_*c*_ are retained. **(b)** Daily new infections for increasing values of the cutoff distance. **(c)** Spatial visualization of the daily new infections using a cutoff *d*_*c*_ = 200 km. See also Movie S3.

### Agreement with 1D analysis of a wave

The results from the previous sections suggest the apparition of a wavefront when transmissions are short range. To investigate this phenomenon, we consider a simplified model of communities lying on a two-dimensional square lattice. Each community index is replaced by coordinates (*i, j*) ∈ ⟦1, 2^*n*^⟧ × ⟦1, 2^*m*^⟧. In particular, we consider that only individuals from neighboring communities interact together. We rewrite equation (1):

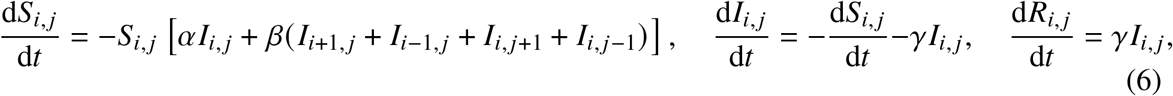

where *α* (respectively *β*) is the intra-community (resp. inter-community) transmission rate.

After rescaling the time and space variables, and defining the rescaled recovery rate 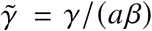, we look for wave solutions in the continuum limit by introducing the shape functions 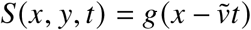 and 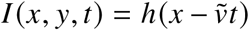, where 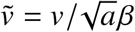 represents the velocity of the wave in rescaled time and space (see SI). The shape functions satisfy the ODE:

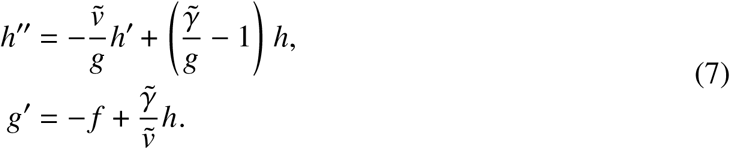

We find that the velocity of the traveling wave is bounded from below (SI):

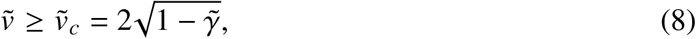

which is in agreement with previous reports [18, 38] and with results from marginal-stability analysis [21, 39, 40]. Interestingly, by an independant argument, we also established (see SI) that 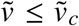. Therefore the traveling wave must move at velocity 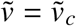. This indicates that the SIR dynamics in equation (6) falls into the Fisher-Kolmogorov-Petrovsky-Piscunov universality class, resulting in pulled waves [41–44]. Although we have taken the continuum limit of a nearest-neighbor model, this analysis is also valid for any finite-range infection matrix with the appropriate rescaling of variables.

We performed simulations of the dynamics given by equation (6) on a square lattice with a varying aspect ratio (Figure 4**a**). As expected, there is a front of new infections, moving from west to east as time progresses. A timelapse of a traveling front of infected individuals with = *β* = *y* = 0.1 is shown in Figure 4**b**. The wave position increases asymptotically linearly with time, but the velocity 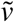 of the wave varies with 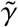 (Figure 4**c**). The profiles obtained are in agreement with the shape functions obtained by solving equation (7), as shown in Figure 4**d**.

**Figure 4.**
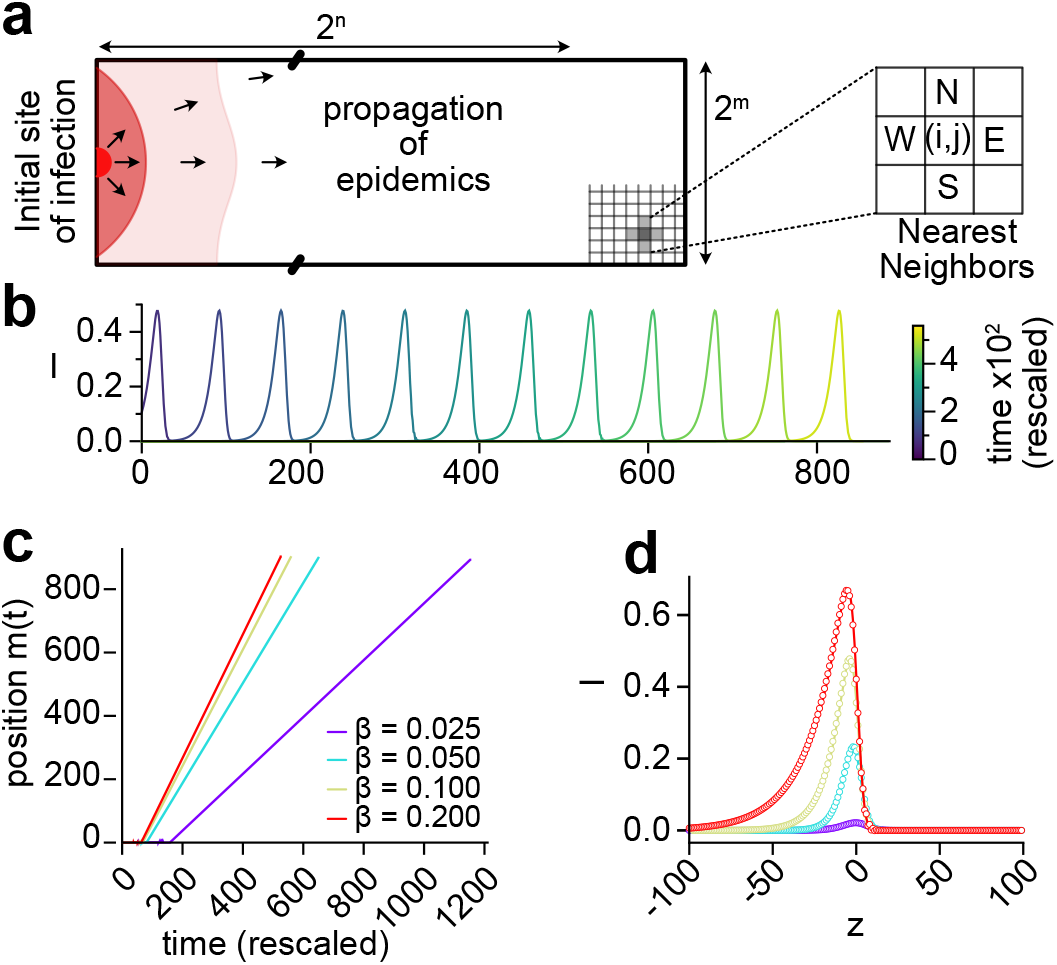
Existence of a wave with nearest-neighbors-only interactions. **(a)** Equation (6) is solved on a square lattice of 2^*n*^ × 2^*m*^ sites. **(b)** Traveling front of infected individuals moving along the direction (left to right). We took *α* = *β* = *γ* = 0.1, so that the rescaled recovery rate is 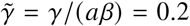. **(c)** The dynamics simulated for different values of *α*. The po sition *m*(*t*) (*S*(*t, m* (*t*)) = (1 + *S*_∞_)/2) of the wave is asymptotically linear in time, hence a constant velocity 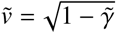. **(d)** The simulated wave profiles (symbols) are in agreement with the profiles predicted by equation (7) (solid lines). The corresponding S,I,R profiles are shown in Figure S2. A simulation with *β* = 0.1 d^−1^ is shown in Movie S4.

### Properties of infectivity matrices

The daily infectivity matrices constructed from the SafeGraph mobility data can be viewed as elements of a random-matrix ensemble. Remarkably, matrix elements seem to be distributed according to the law *β*_*ij*_ = ⟨*β*_*ij*_⟩*e*^*ξ*^, where ⟨*β*_*ij*_⟩ is the mean infectivity matrix and *ξ* ≡ *N*(0, 1) is a centered reduced Gaussian variable (Figure S3). The probability density of the eigenvalues is also shown in Figure S3. A connection can be made with random matrix theory (RMT) [45–47], initially introduced by E. Wigner to model the spectra of the nuclei of heavy atoms, where the interactions between many nucleons are assumed to be drawn from a random ensemble. In RMT, random matrices are classified according to their symmetry, or according to their corresponding level statistics (namely, the probability density of the spacing between consecutive eigenvalues) that exhibit different degrees of level repulsion [45]. In particular, the Wigner-Dyson (WD) statistics is typical of the Gaussian Orthogonal Ensemble (in which eigenvalues “interact”), while the Poisson statistics is typical of a matrix with independent eigenvalues. The level-spacing statistics of the infectivity matrices is shown in Figure 5**a**. We found that they were time-independent. Yet interestingly, the level statistics interpolates between the WD statistics and the Poisson statistics [48]. As shown earlier, removing links between communities according to their geographical distance results in a slower spread and a smaller epidemic size. Concomitantly, the level spacing distribution converges toward a Poisson statistics (Figure 5**c**). The crossover from WD (entropy *S* = 0.7169) to Poisson (*S* = 1) distribution as links between communities are successively removed suggests an isolation policy that can lead to an effective reduction of the epidemic size (SI). As alternative mitigation strategies, we choose to induce the transition toward a Poisson distribution by decimating links between communities according to their “nominal” distance or the “edge-betweenness” centrality [49–51] (Figure 5**b**). Figure 5**d** shows how the level spacing distribution converges toward a Poisson statistics when the nominal distance threshold is lowered. We find the edge-betweenness centrality to be more efficient in decreasing the epidemic size. This is because decimation according to edge-betweenness first targets links with the largest transmission rates. One could also consider a moderate policy: instead of eliminating links completely, one could impose constraints on the flux of individuals that are allowed to commute via central pre-determined links.

**Figure 5.**
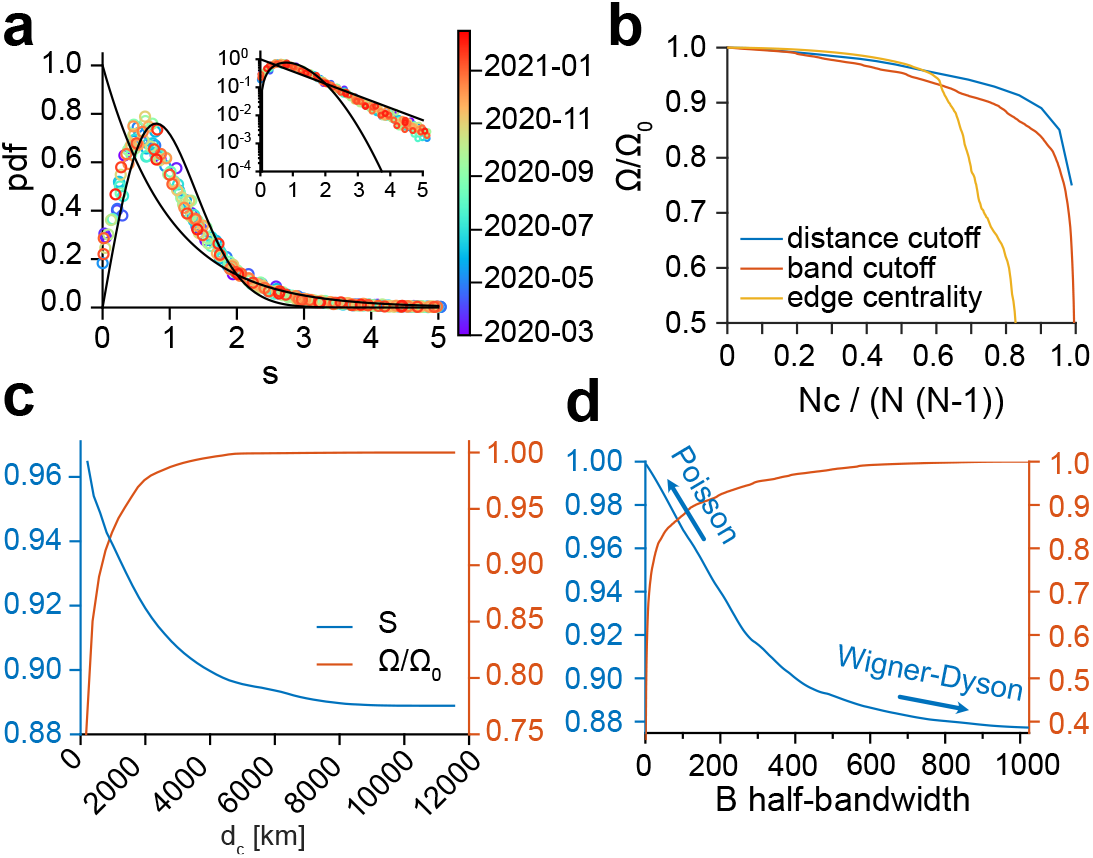
Random-matrix properties and mitigation strategies. **(a)** Level spacing distribution (unfolded [45, 52–54]) of infectivity matrices between 2020-03-01 and 2021-02-15 (pooled by 7-day windows). The solid lines denote the Wigner-Dyson and the Poisson statistics. The level distribution is shown in Figure S3. **(b)** The epidemic size Ω/ Ω_0_ versus the relative number of cuts *N*_*c*_/*N*/(*N* − 1) for the 3 mitigation methods: distance cutoff, bandwidth, and edge-betweenness centrality. Ω_0_ is the epidemic size for *N*_*c*_ = 0. Level-spacing entropy *S* and epidemic size as a function of **(c)** the cutoff distance *d*_*c*_ in the range 170-12000 km, and **(d)** the half-bandwidth 1≤*B <* 1023.

## Discussion

Biological systems are inherently complex and it can be a challenge to characterize them by a small number of parameters. In the case of epidemics, the huge number of parameters (for example, intercommunity spread involves *N*^2^ transmission rates for *N* communities) can obscure the salient mechanisms and make it difficult for policy makers to find efficient interventions. Reducing the dimension of a model, when possible, is therefore of great value. In this article, we have proposed a spatial SIR model with an infectivity matrix based on the local travel patterns. Despite its simplicity (there is only one global parameter to be adjusted), we find that our model is able to capture the spatial spread of the SARS-CoV-2 epidemic, with a delocalized multicenter spreading caused by long-distance travelers bringing infection into far regions, then becoming secondary centers of infection. The time evolution of the interaction frequency reflects the shelter-in-place policies that were implemented by various USA states in the early stages of the SARS-CoV-2 pandemic. In fact, the rich diversity of human responses could be summarized by a simplified model for the interaction frequency, whose asymptotes represent the values “before lockdown” and “after lockdown”, with few assumptions about the infection process. The individual variances between people and communities turn out to self-average, giving a clear picture of the spread.

If a model with relatively few parameters describes the observations, one can assume that it also describes the situation when these parameters are changed by an intervention. Therefore, we suggest such a model could be used as the basis for efficient policies—or at least reliably to estimate the consequences of adopting policies. As an example, we have shown the hypothetical effect of an alternative to the shelter-in-place policy in the case of the SARS-CoV-2 pandemic. Specifically, we have investigated a travel restriction policy in which individuals can only move within an area of fixed radius centered around their residence. By contrast to the lockdown policy, we find that infections spread through a well-defined wave front, traveling with a certain velocity. This scenario might be preferable since it gives time to communities and public health infrastructures to prepare for the onset of the epidemics, while still “flattening the curve” of new infections (Figure 3**b**). We have also provided both an analytical and numerical analysis elucidating the mechanism of formation of a wavefront. In particular, we give the velocity and the shape of the wavefront for an epidemic spreading through nearest neighbors interactions.

Although SafeGraph mobility data [36] provide a realistic picture of people movement between communities, one might ask what is the sensitivity of the results to the infectivity matrix derived from the mobility fluxes. We thus considered a model with uniform transmission rates among communities, namely *β*_*ab*_ = *β*, which suppresses spatial effects. In particular, this model leads to the natural variables *v*_*a*_ (see SI) to be uniform: *v*_*a*_(*t*) = *v*^UN^(*t*). Surprisingly, carrying out the same fit to the reported cases (Figure 1**a-b**) was only marginally inferior to the fit carried out with the SafeGraph mobility data (see Figure S1**b**). By contrast, carrying out the same fit with the infectivity matrix derived from SafeGraph mobility data but with long-range interactions turned down resulted in a significantly different dynamics (see Figure S1**c**). There are several implications of this result. (1) Although changes in the structure of the infectivity matrix can lead to drastically different dynamics (Figures 3 and 5 and Figure S1**c**), it appears that the infectivity matrix derived from SafeGraph fluxes falls in the same “universality class” as the uniform model. This suggests that long-range movements (e.g. air traffic) played a prevalent role in the spread of SARS-CoV-2. (2) Our choice to reduce the complexity of the model to only one fitting parameter might not be adequate to discriminate between models falling into the same “universality class”. Instead of fitting only one scalar *p*(*t*) at each time point, we also investigated the possibility of fitting the *N*^2^ transmission rates *β*_*ab*_(*t*) minimizing the errors with reported cases (see Material and Methods and Figure S4). Although this latter approach is clearly prone to overfitting, it shows that there exists a parametrization of the model which reproduces very closely reported cases. We anticipate that the proper number of fitting parameters lies in between those two extreme scenarios.

During the course of our research, an epidemiological study bearing similarities with our approach was published [55]. In that study, the authors developed a county-resolved metapopulation model describing the spread of SARS-CoV-2 in the USA, informed with mobility data from SafeGraph. Instead of calibrating the model by fitting the country-wide number of reported cases as in the present study, the authors fitted their model to reproduce reported cases of COVID-19 on a per-county basis. Furthermore, the model required fitting many time-dependent parameters on a per-county basis, including per-county transmission rates, making the fitting procedure very high dimensional. Finally, the dynamics of the disease spread introduced differs from equation (1) since *S, I* and *R* compartments were introduced for each commute channel *i* → *j* rather than for each community. Altogether, the complexity of that model makes it less amenable to analytical study.

In conclusion, we used a simple model of intercommunity spread of an infectious disease to show the transition between different regimes of epidemic progression. Because of the complexity of the infection process (e.g., variations in individuals’ responses, mutations, etc.) and of human behavior, we are still far from a global forecasting system able to predict the spread of different infectious agents throughout the world. As with weather forecasting, observables must be measured in real time in order to inform complex models to yield short-term forecasts. One salient feature in our approach is showing how such widespread measurements (namely, the mobility data) can be integrated in a model describing the spread of an infectious agent, and showing what types of predictions can be obtained. It is a step toward more predictive epidemiology models, grounded in measurable quantities.

## Material and Methods

### Data availability

Data and scripts used in this study are available at the GitHub repository github.com/czbiohub/epidemiology_flux_model. Except the raw mobility data, which belongs to SafeGraph.

### Mobility data

The mobility data was obtained from [SafeGraph](https://www.safegraph.com/), a company that aggregates anonymized location data from numerous applications in order to provide insights about physical places, via the SafeGraph Community. To enhance privacy, SafeGraph excludes census block group (CBG) information if fewer than two devices visited an establishment in a month from a given census block group. In this manuscript, we use data extracted from the “Social Distancing Metrics” dataset [36], with dates between February 2020 and February 2021. SafeGraph has stopped sharing mobility data under this format. However the matrices of fluxes between our coarse-grained communities can be found in the ‘data’ folder in our github repository. We obtained the coarse-grained communities by running a *K*-means clustering algorithm to group the 220, 333 CBGs into 2^10^ communities. We used the implementation from [Scikit Learn](https://scikit-learn.org/stable/modules/clustering.html). We then ran a hierarchical clustering algorithm to re-index communities so that communities close in space had close indices, as shown in Figure 1. We used the [linkage](https://docs.scipy.org/doc/scipy/reference/generated/scipy.cluster.hierarchy.linkage.html) function from SciPy. We then computed for every day the “flux matrix” where each entry *f*_*ab*_ represents the number of cell phone whose residence CBG belongs to community *a* which visited a CBG belonging to community *b*. The average flux matrix was constructed by averaging all the daily flux matrices. We used population counts for CBGs in agreement with the United States Census Bureau’s and available in the [SafeGraph Open Census Data](https://docs.safegraph.com/docs/open-census-data) (file “cbg_b01.csv”, column “B01001e1”). We checked that the population counts from the United States Census Bureau were approximately proportional to the residential mobile-phone counts, therefore validating mobile tracking as a proxy for actual population.

### Reports on SARS-CoV-2 infections

In order to fit our model, we used USA cases of COVID-19 reported by the Center for Systems Science and Engineering (CSSE) at Johns Hopkins University [37] which can be accessed at the GitHub repository CSSEGISandData/COVID-19. New cases of COVID-19 reported by the CSSE were mapped to the closest mobility-data-derived community based on their latitude and longitude. We therefore obtained the time evolution of SARS-CoV-2 infections through the mobility-data derived communities. Integer absolute new cases were converted into relative population fractions using the community population counts obtained from the United States Census Bureau.

### Model fit

#### Daily scales

Here we consider the SIR dynamics (see Supplementary Information):

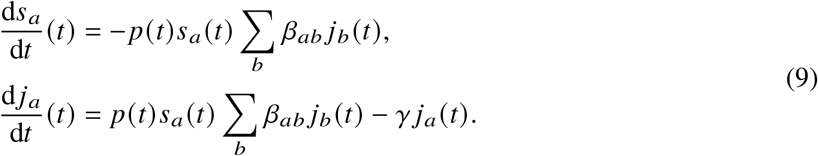

We also define the chi-square at each time *t*:

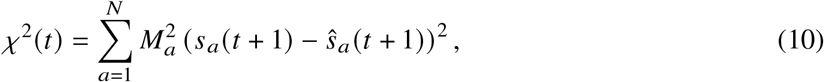

where *M*_*a*_ is the population at location *a* and (1 − *ŝ*_*a*_)*M*_*a*_ is the number of reported cases at location *a*. Assuming that *p*(*t* − 1), *p*(*t* − 2), …, *p*(0) have been previously evaluated, the dynamics (*s*_*a*_, *j*_*a*_) is determined up to time *t*, and we have:

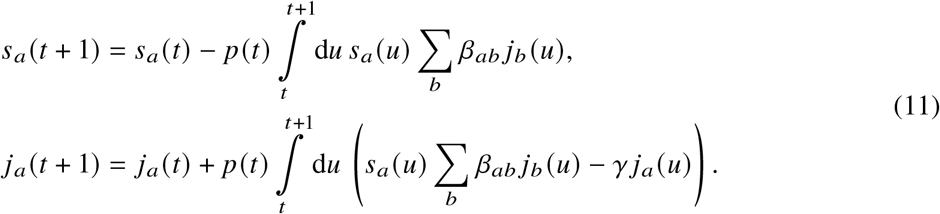

To obtain the scale *p*(*t*), we solve for:

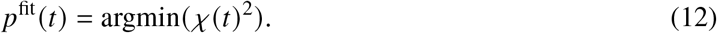

#### Simplified model

We look for a simplified model in which the scales have a functional form close to a ramp function:

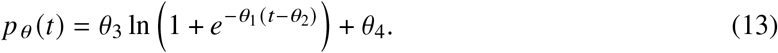

We first set *θ* by fitting this function to the daily scales:

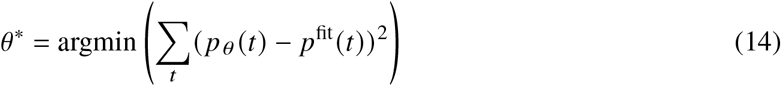

Then we adjust the slope of the ramp in order to minimize the error with reported cases. In particular, we set 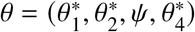, and we solve:

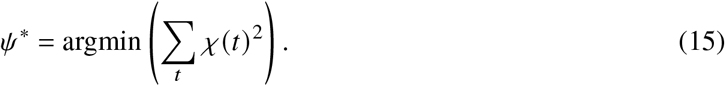

We thus define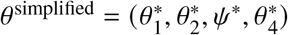, and the model for simplified scales is:

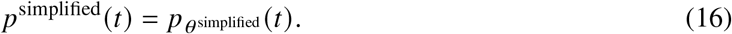

#### Daily infectivity matrices

We also considered another approach, consisting of fitting the *N*^2^ transmission rates *β*_*ab*_(*t*) at every time *t*. In particular, we solve:

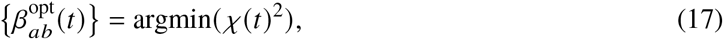

subject to the constraints *β*_*ab*_ *≥* 0. We show the results of this fit in Figure S4, however his approach is prone to overfitting since there is *N*^2^ fitting parameters and only *N* data point at each time *t*.

### Simulations with nearest-neighbors-only interactions

The curves shown in Figure 4**b-c** and the symbols shown in Figure 4**d** were obtained by integrating equation (6).We used the function solve_ivp from SciPy with the “DOP853” integration method. At *t* = 0, we considered *S* = 1 everywhere except at the sites of coordinates (0, 2^*m*−1^ − 1) and (0, 2^*m*−1^) (see Figure 4**a**), where we set *S* = 0 and *I* = 1. The Laplacian was computed using the 9-point stencil 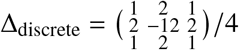. We considered periodic boundary conditions along the *y* direction and Dirichlet boundary conditions along the *x* direction.

## Data Availability

All data produced are available online at https://github.com/czbiohub/epidemiology_flux_model and upon reasonable request to the authors.

## Acknowledgements

The authors acknowledge the generous support of the Chan Zuckerberg Initiative and Chan Zuckerberg Biohub. The authors also thank Mark Rychnovsky for useful conversations on random matrix theory in the early stages of this project, and Naomi Rankin for her help with a preliminary version of the lattice simulations. D.Y. acknowledges support by MINECO (Spain) through Grant No. PGC2018-094684-B-C21, partially funded by the European Regional Development Fund (FEDER). We also are grateful to the anonymous reviewers of *Scientific reports* who asked deep questions, that helped us to better understand our results.

## Supplementary Information

### 1 Supplementary figures

**Figure S1.**
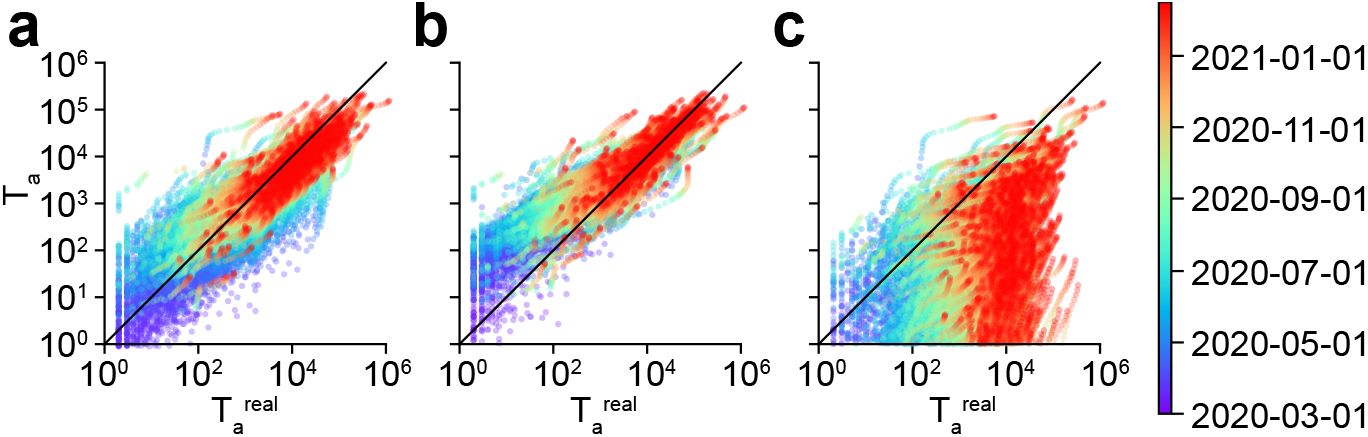
Supplementary figure to Figure 2 of the main text. Comparison of the model predictions with reported values for **(a)** the infectivity matrix derived from SafeGraph fluxes, **(b)** the uniform infectivity matrix, and **(c)** the infectivity matrix derived from SafeGraph fluxes truncated to geographical distances *d*_*c*_ *<* 400 km (breaking of universality class). Local epidemic sizes *T*_*a*_ are shown. One symbol is associated to one given community and one given day.

**Figure S2.**
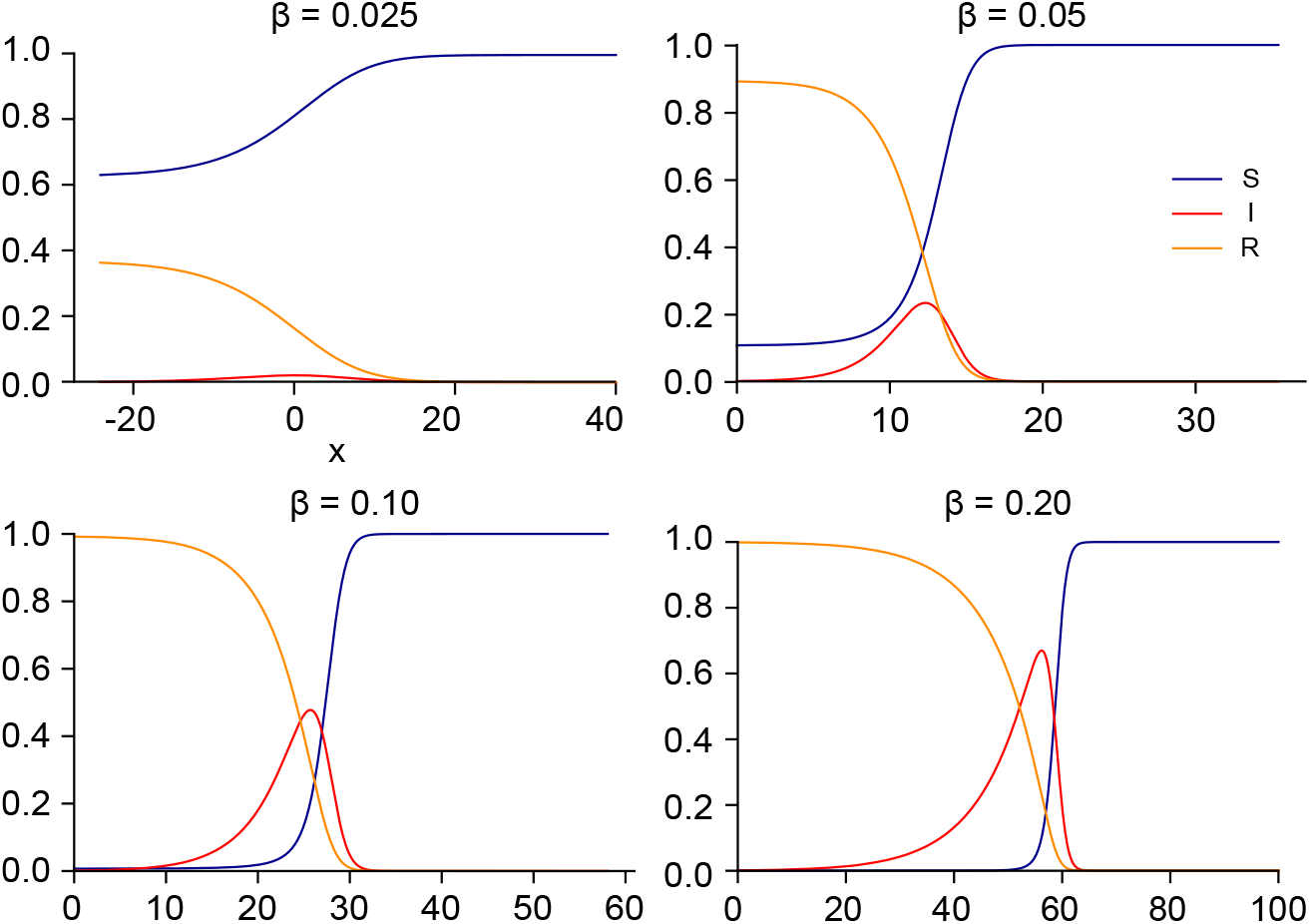
Supplementary figure to Figure 4 of the main text. Wave profiles for the values of *β* shown in Figure 4**d-e**.

**Figure S3.**
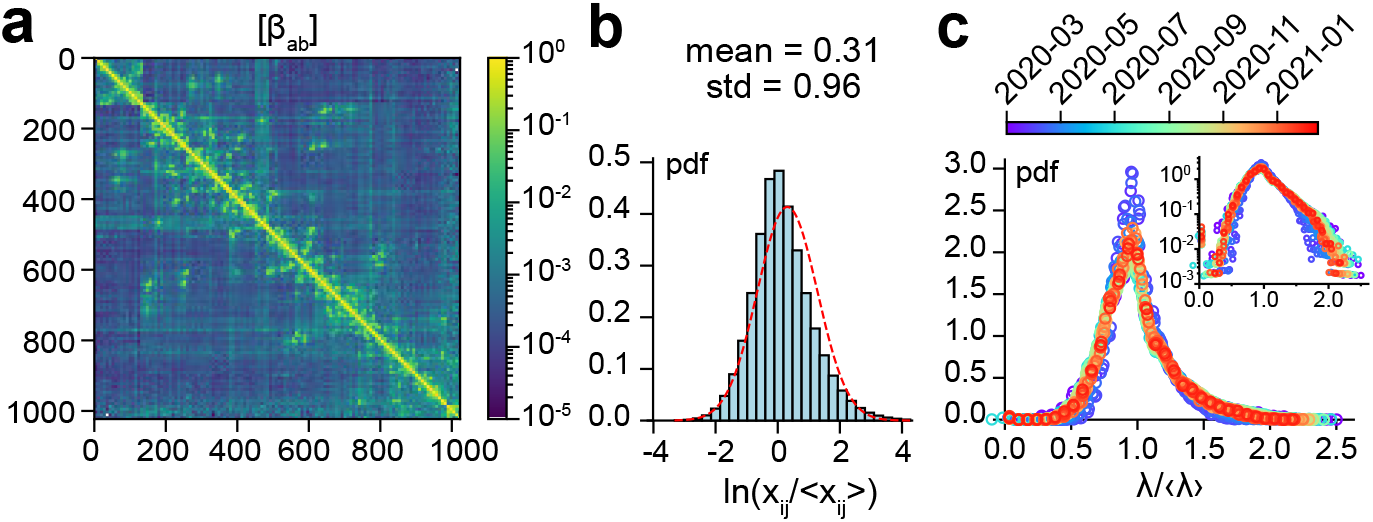
Supplementary figure to Figure 5 of the main text. **(a)** Mean infectivity matrix (between 2020-03-01 and 2021-02-15). Entries were pooled in squares of size 8 × 8. The maximum is shown. **(b)** The noise distribution suggests a log-normal distribution of each entry around its mean. **(c)** Eigenvalue distribution of infectivity matrices as a function of time (pooled by 7-day windows).

**Figure S4.**
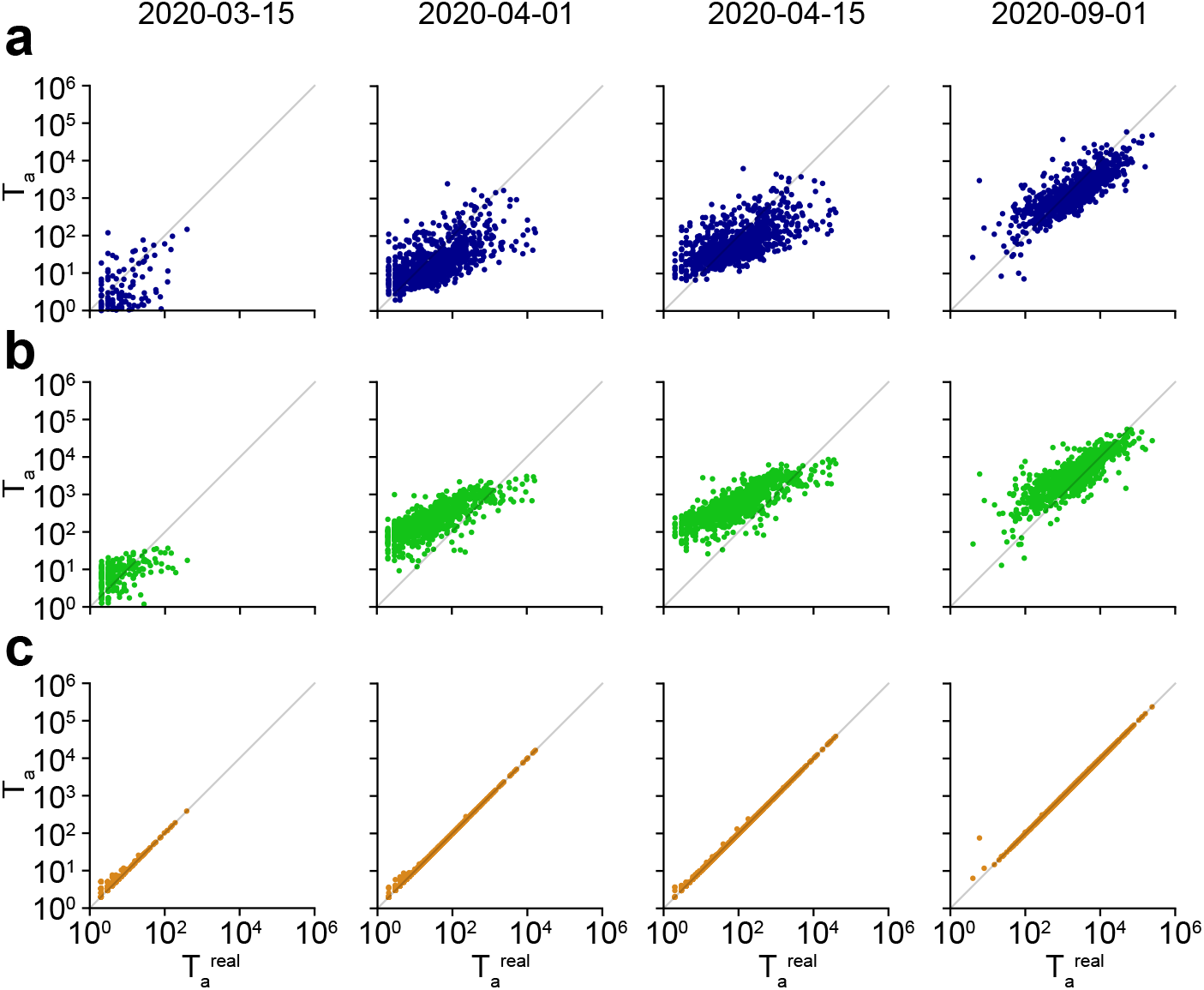
Comparison of instantaneous model predictions with reported values at different dates. **(a)** Scale fitting procedure with infectivity matrix derived from SafeGraph mobility data. **(b)** Scale fitting procedure with uniform infectivity. **(c)** Fitting procedure with *N*^2^ transmission rates *β*_*ab*_ (*t*).

### 2 Formal solution to the spatial SIR

#### 2.1 Matrix SIR

We consider *N* communities affected by a growing epidemic. The spreading of the epidemic is described by the multi-compartment SIR model. Every community *a* is characterized by the number of susceptible individuals *S*_*a*_, the infected individuals *I*_*a*_ and the number of recovered individuals *R*_*a*_. The dynamics for *S*_*a*_, *I*_*a*_ and *R*_*a*_ with *a* ∈ ⟦1, *N*⟧ is described by the equations

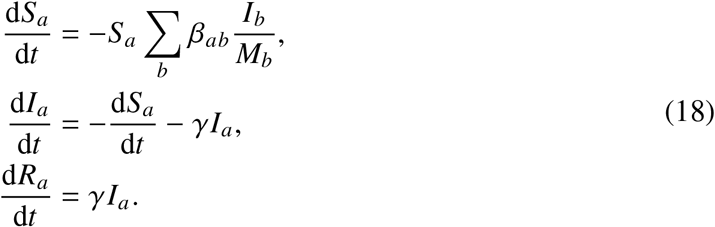

We assume that the population in each community is fixed, i.e.

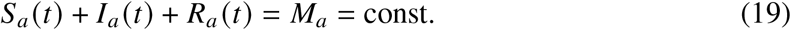

The boundary conditions for equation (18) are:

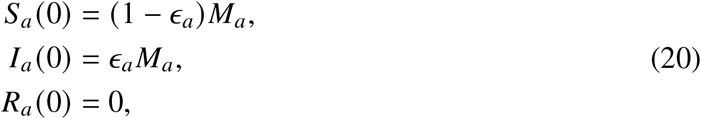

where the parameter *ϵ*_*a*_ describes the initial infection (the initial number of infections per per person in the given community).

We introduce the rescaled variables: *s*_*a*_ = *S*_*a*_/*M*_*a*_, *j*_*a*_ = *I*_*a*_/*M*_*a*_ and *r*_*a*_ = *R*_*a*_/*M*_*a*_. Then equations (18) and (20) become

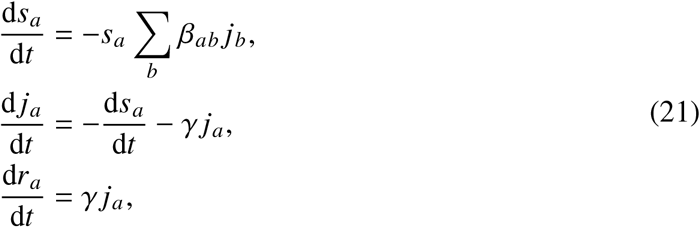

with

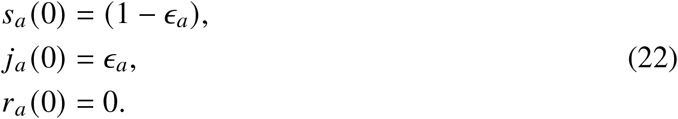

#### 2.2 Solution of matrix SIR equations: the canonical basis

In order to solve equations (21) and (22), we define:

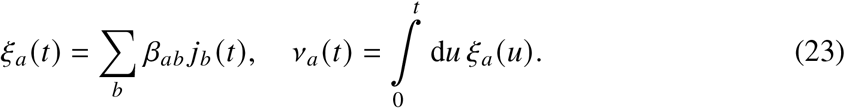

Substituting equation (23) in equation (21), we solve for *s*_*a*_ and obtain:

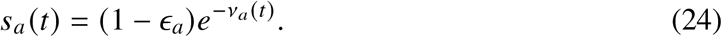

We also note that *r*_*a*_ (*t*) can be written as:

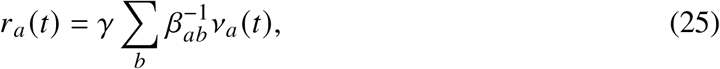

where 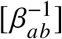 denotes the inverse matrix such that 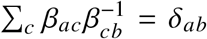. We therefore obtain the parametrization:

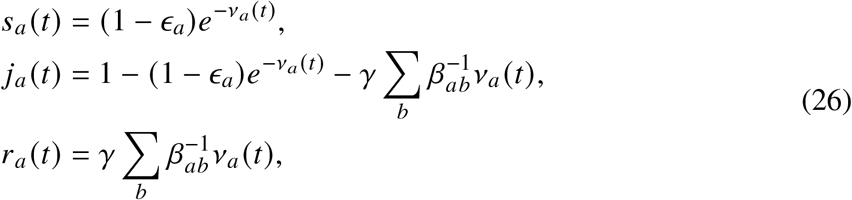

as a function of *v*_*a*_ (*t*). Note that when the infectivity matrix is diagonal, namely *β*_*ab*_ = *βδ*_*ab*_, we recover the parametrization from reference [4]:

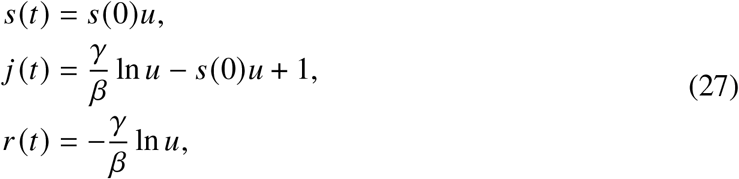

with *u*(*t*) = *e*^−*v*(*t*)^.

The dynamics is determined by the functions *v*_*a*_(*t*). Starting from equation (21), we multiply the equations by *β*_*ab*_ and sum, obtaining

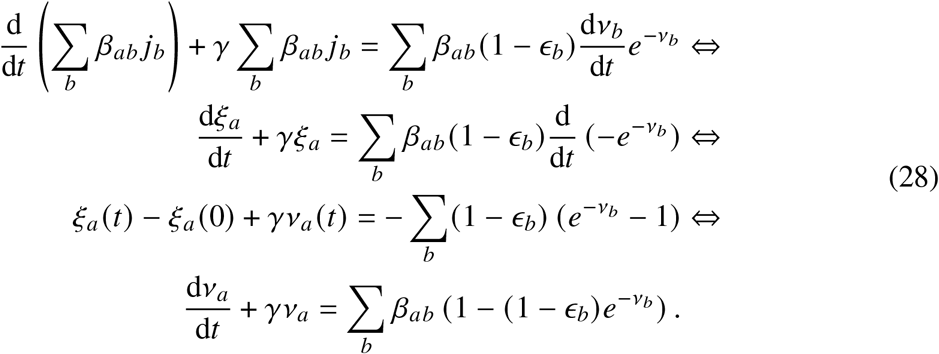

Equation (28) can be solved numerically. In the *t* → ∞ limit, we obtain:

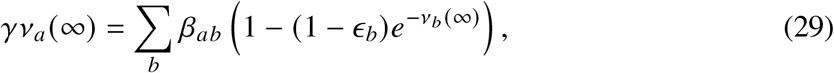

which is the same as setting *j*_*a*_ = 0 in equation (26). Equation (29) can be solved iteratively. The size of the epidemic is given by:

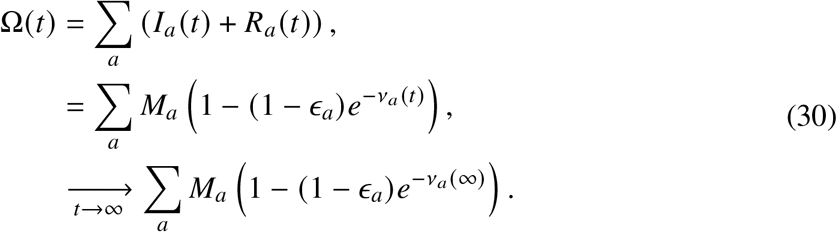

#### 2.3 Solution of matrix SIR equations: SVD basis

A solution can also be written using the singular value decomposition of the infectivity matrix *β*:

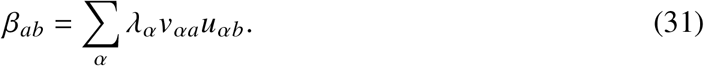

For a given vector **x**, we use the following notation: 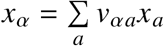. From equation (28) we thus obtain:

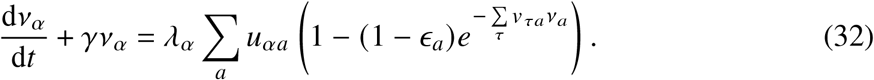

### 3 Initial stage of an epidemic

We again start from equation (21), which we linearize around the initial state 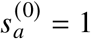, 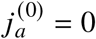, and 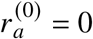. To first order **j** and **r** satisfy the ODE:

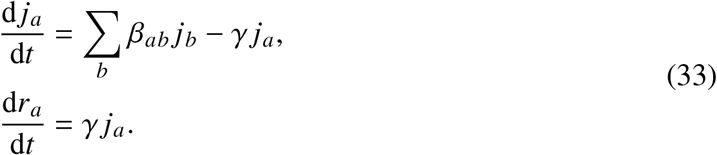

[*β*_*ab*_] is a real square positive matrix. According to the Perron-Frobenius theorem, there exists the maximal eigenvalue *λ*_*ω*_ > 0, such that any other eigenvalue *λ*_*α*_ < *λ*_*ω*_, and the associated eigenvector **v**_*ω*_ is positive. At large times, we have:

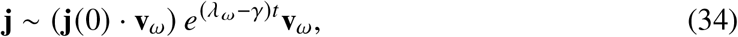

where (**A** · **B**) denotes a scalar product. The epidemic grows only if the basic reproduction number 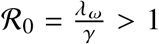. Furthermore, if the infectivity matrix can be factorized as *β*_*ab*_ = *f*_*a*_*g*_*b*_, then *β* is of rank 1 and *λ*_*ω*_ is the only non-zero eigenvalue. We then have 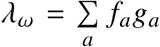 and **v**_*ω*_ = **f**.

### 4 Construction of the infectivity matrix from mobility data

The mobility data obtained from SafeGraph allows us to construct a pseudo-flux matrix in which each entry *f*_*ab*_ represents the number of individuals from community *a* visiting community *b* per day. Let us now consider one community *a*, having *S*_*a*_ susceptible individuals and *I*_*a*_ infected individuals. The variation in susceptible individuals due to new infections during the time interval Δ*t* has the form:

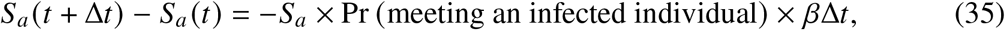

where *β* Δ*t* represents the probability to get infected when meeting an infected individual. We now list the different contributions.

#### 4.1 Intra-community contributions

The contributions coming from infected individuals in the same community are:

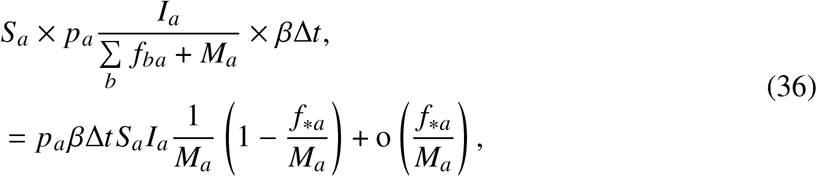

where we have introduced *f*_\**a*_ = ∑_*b*_ *f*_*ab*_, and the parameter *p*_*a*_ representing the frequency with which an individual is interacting with other individuals in community *a*. Here *I*_*a*_/(*f*_\**a*_ + *M*_*a*_) is the probability to meet an infected individual when interacting with an individual in the community.

#### 4.2 Inter-community contributions: incoming visitors

The contributions to the infections in community *a* from infected individuals visiting from another community *b* are:

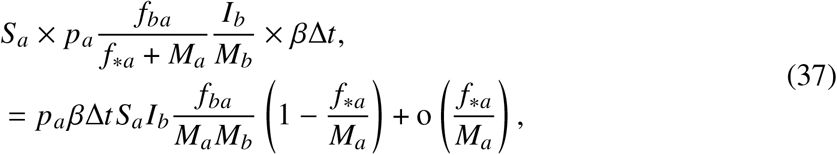

where we have assumed that *f*_*ba*_*I*_*b*_/*M*_*b*_ is the number of infected visitors from *b*.

#### 4.3 Inter-community contributions: returning natives

The contributions coming from individuals from community *a* infected while visiting another community *b* are:

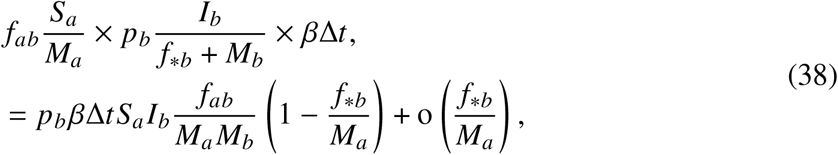

where we have assumed that *f*_*ab*_*S*_*a*_/*M*_*a*_ is the number of susceptible visitors from *a* visiting community *b*.

Neglecting the *f*_∗*a*_*/M*_*a*_ terms, we obtain after adding all contributions:

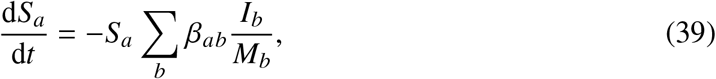

with:

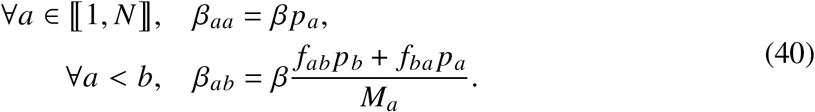

Note that in general, the matrix [*β*_*ab*_] is not symmetric. In the manuscript, we make the assumption that the interaction frequency is the same in all communities, namely *p*_*a*_ = *p*.

### 5 Spreading with a wave of infection

#### 5.1 ODE for the wave profile

We consider the SIR model on a 2d lattice, with infections limited to nearest-neighbors, and with uniform population *M*_*a*_ = *M*. For the sake of simplicity, we will consider the rescaled *S, I, R* variables with *S* +*I* + *R* = *M* = 1. We write:

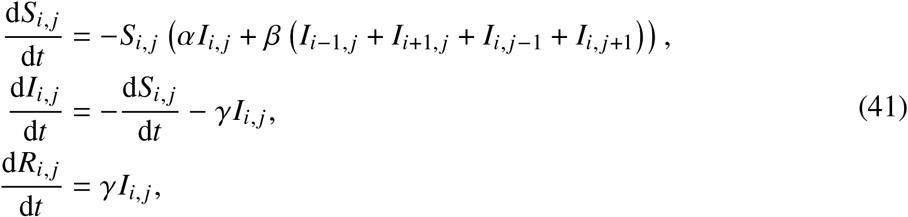

where *i* and *j* denote the indices along the first and second dimensions. We introduce the discrete laplacian:

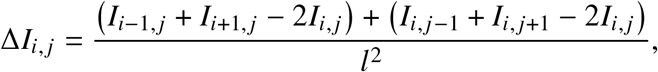

where *l* is the lattice spacing. Equation (41) becomes:

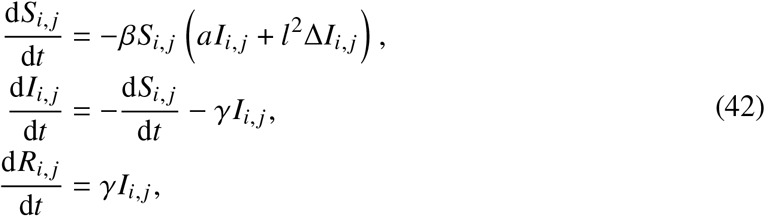

where *a* = 4 + *α*/*β*. In this study, we will consider *α* = *β*, so *a* = 5. In the continuum we have:

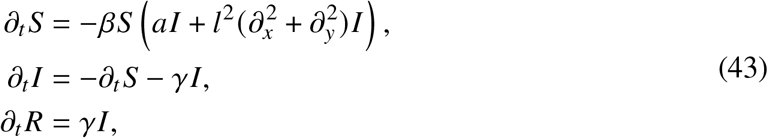

Limiting ourselves to solutions which only depend on one space variable, namely *S*(*t, x, y*) = *S*(*t, x*), and after rescaling the time variable, *t* ← *t*(*aβ*)^−1^, the space variable, *x* ← *xla*^−1/2^, and the recovery rate, *γ* ← (*αβ*)*γ*, we obtain:

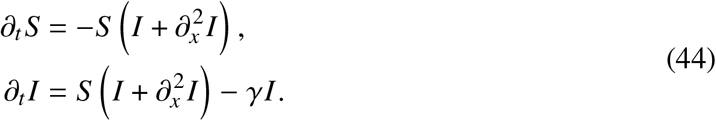

We are interested in the traveling front solutions, namely *S*(*x, t*) = *g*(*x* −*vt*) and *I*(*x, t*), = *h*(*x* − *vt*), where *v* is the velocity of the traveling waves. Here, *g*(*z*) and *h*(*z*) are the shape functions for the propagating front. Making those substituations in equation (44), we obtain the ODE:

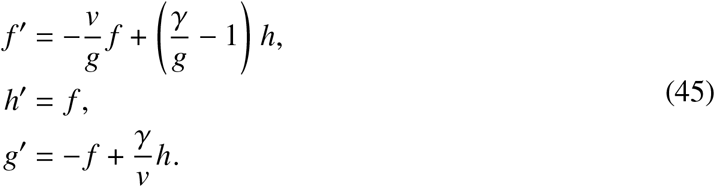

#### 5.2 Velocity selection

The solution to equation (45) specifies the shape functions *g* and *h* for a traveling front solution with velocity *v*. We now characterize what are the admissible velocities. We start by computing the jacobian of the function on the right-handside of equation (45):

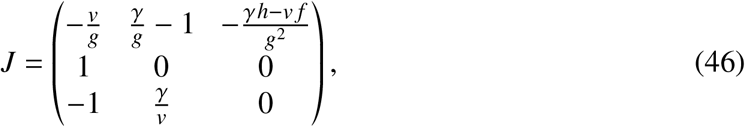

and its characteristic polynomial:

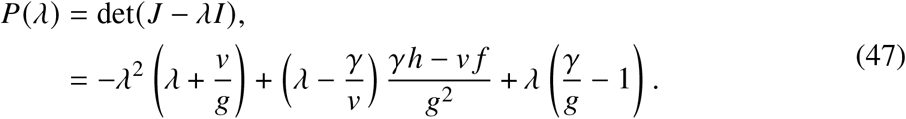

The fixed points of the ODE in equation (45) are of the form (0, 0, *g*), with *g* ∈ [0, 1]. At those points, there is one zero eigenvalue, and two eigenvalues satisfying the equation:

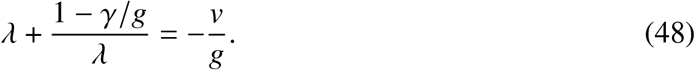

The fraction of susceptible individuals *S*(*x,t*), is a decreasing function of *t*. Thus it forbids any oscillatory behavior in the shape function *g*(*z*). Therefore the two eigenvalues must be real, which results in the condition:

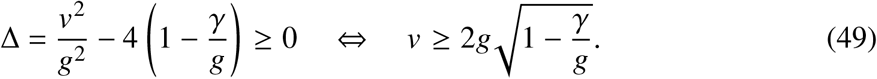

The function in the right-handside of the inequality is an increasing function of *g*. Furthermore, the fixed point (0, 0, 1) must belong to the front solution since it corresponds to the unstable initial condition with only susceptible individuals. Therefore, enforcing that all admissible fixed points have real eigenvalues yields the condition:

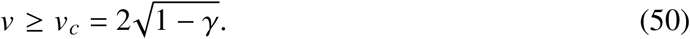

#### 5.3 Upper bound on the velocity

Since *S* ≤ 1, by retracing the steps from equation (41) to equation (44) we have that:

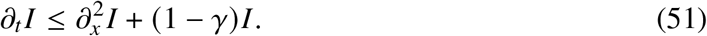

Let us define *Ĩ*, which is a solution to

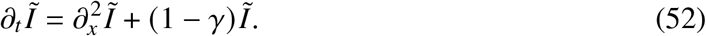

Starting from the same initial condition, *I*(0, *x*) = *Ĩ*(0, *x*), we must have at all times:

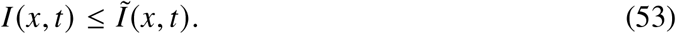

The function *ϕ*(*x*) = (1 − *γ*)*x* satisfies the condition [41] *ϕ*′ (*x*) ≤ *ϕ*′(0) for all *x* ∈ [0, 1], therefore equation (52) falls into the Fisher-Kolmogorov-Petrovsky-Piscunov (FKPP) universality class [41–44]. In particular, for a step initial condition, *Ĩ*(*t, x*) evolves into a moving front with velocity 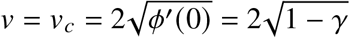.

Because of the inequality in equation (53), any moving front *I* must move at a velocity smaller than the velocity of the moving front *Ĩ*. Otherwise, the *I* front would eventually pass the *Ĩ* front, which violates equation (53). Therefore, the velocity of the front *I* must satisfy:

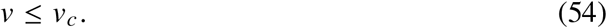

**Figure S5.**
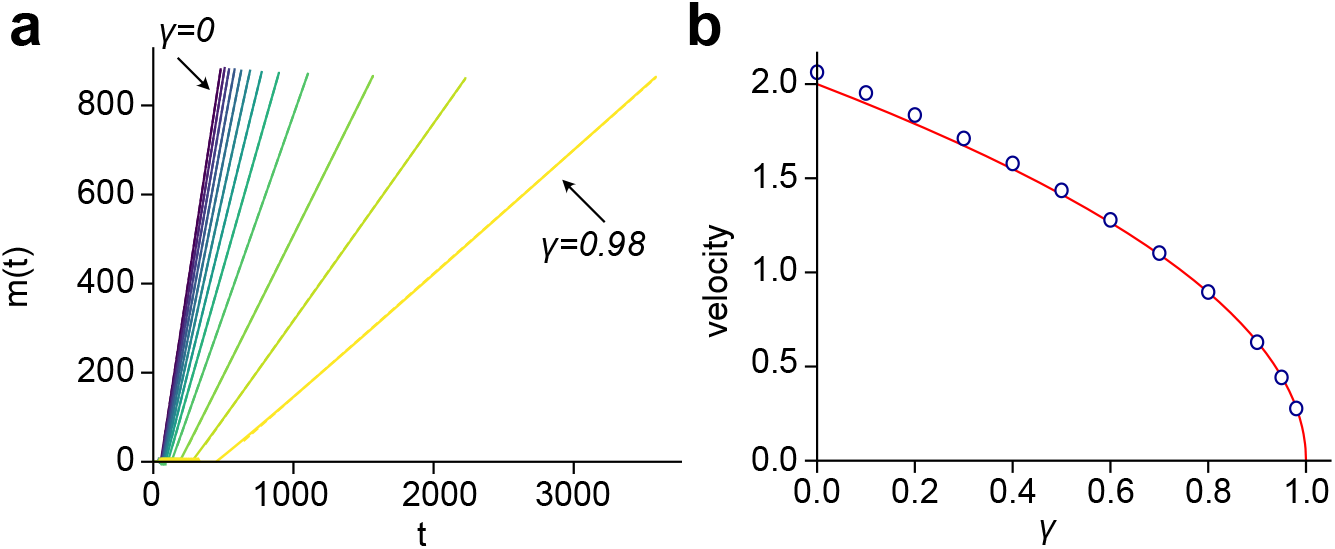
Traveling wave solutions to equation (44). **(a)**. Position of the moving front as a function of time, *m*(*t*), for values of *γ* = 0, 0.1, 0.2, 0.3, 0.4, 0.5, 0.6, 0.7, 0.8, 0.9, 0.95, 0.98. **(b)** Fitted velocities collapse on the theoretical *v* = *v*_*c*_.

#### 5.4 Pulled wave

Combining equations (50) and (54), we obtain that any moving front resulting from equation (44) moves at velocity 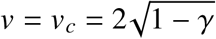. Therefore it falls in the FKPP universality class and it is a pulled wave.

We have solved numerically equation (44) for several values of *γ*, starting from the initial condition: *S*(0, *x*) = 1, *I*(0, *x*) = 0, ∀*x* > 0 and *S*(0, 0) = 0, *I*(0, 0) = 1. In Figure S5**a**, we show the position of the moving front as a function of time. The position of the moving front, *m*(*t*), was defined such that *S*(*t, m*(*t*)) = (1 + *S*_∞_)/2. Clearly, after a transient regime, the solution evolves into a moving front. We plotted the velocity *v* as a function of *γ* in Figure S5**b**, which agrees with the theoretical prediction 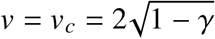.

#### 5.5 Shape of the traveling front

Since a traveling front solution must travel at velocity *v*_*c*_, it follows that the shape is uniquely determined by the recovery rate *γ*. The shape functions *g* and *h* satisfy the ODE:

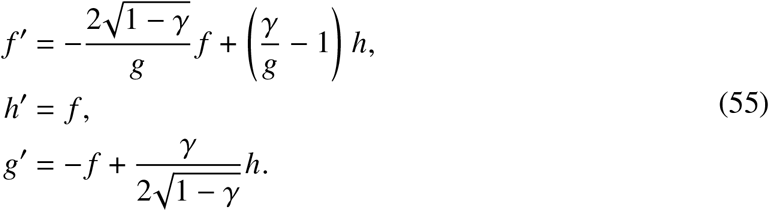

The shape of the traveling front must be such that:

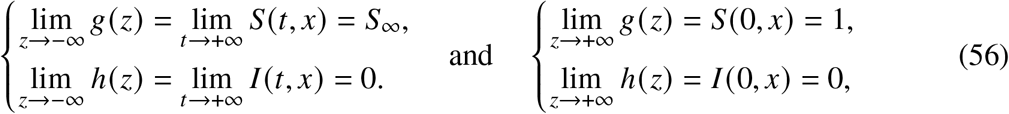

We determine the residual fraction of susceptible individuals, *S*_∞_, using the parametrization in equation (27) with *u* = *e*^−*v*^, and solving:

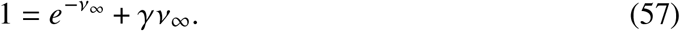

Since the last equation is solved numerically, it is useful to derive two bounds. First, the condition *S*_∞_ > 0 yields the condition *v*_∞_ < *v*^∗∗^ = 1/*γ*. Second, the function *e*^−*x*^ + *γx* − 1 is strictly negative in the interval]0, *v*_∞_[, with a minimum reached at *v*^∗^ = − ln *γ*. Thus we have *v*^∗^ < *v*^∞^ < *v*^∗∗^.

**Figure S6.**
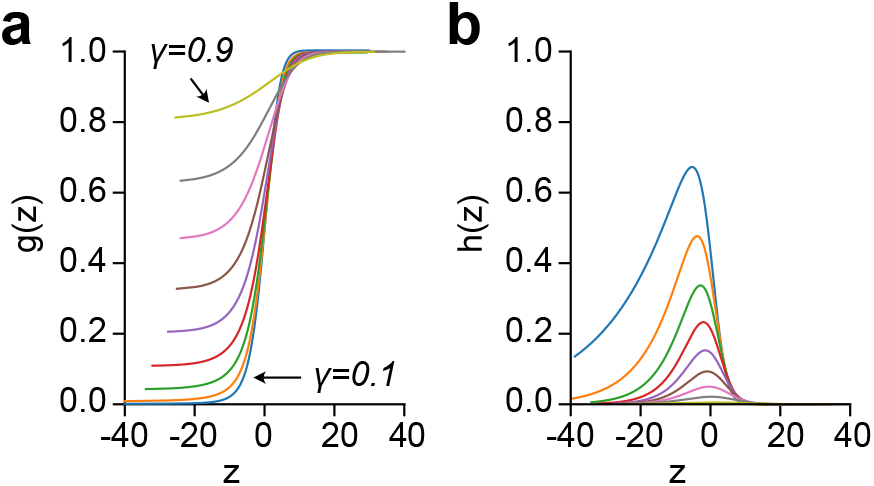
Shape of the SIR traveling waves: **(a)** *g*(*z*) and **(b)** *h*(*z*), obtained by solving equation (55) for values of *γ* = 0, 0.1, 0.2, 0.3, 0.4, 0.5, 0.6, 0.7, 0.8, 0.9.

Following the stability analysis introduced hereabove, (0, 0, *g*) is an unstable fixed point of the trajectory (the jacobian has one positive eigenvalue) as long as *g* < *γ* Therefore, 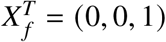 is a stable fixed point of the trajectory (the jacobian has only negative eigenvalues), whereas 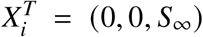 is an unstable fixed point. We can therefore solve the ODE in equation (55) with the initial condition *X* = *X*_*i*_ and let the trajectory converge toward *X* _*f*_. Wave shapes for several values of *γ* are shown in Figure S6.

### 6 Level statistics and connectedness

#### 6.1 The level spacing distribution of the flux matrix

We consider the level statistics of the symmetric matrix *F*_*ab*_ = (*f*_*ab*_ + *f*_*ba*_)/(*M*_*a*_*M*_*b*_), where *f*_*ab*_ is the number of people residing in community *a* visiting community *b* during one day and _*a*_ is the total population of site *a*, see equation (19). The symmetric flux is related to the infectivity matrix through *β*_*ab*_ = *pβF*_*ab*_*M*_*b*_, where the factor *pβ* is estimated by a fit to the SafeGraph mobility data over *K* = 353 days from March 1^st^ 2020 to February 16^th^ 2021 and is given by *pβ* = 0.05 ± 0.02, as is shown in Figure 2**b** of the main text. Following common practices in the treatment of random matrices [45], the diagonal elements 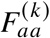 of the *K* matrices of size *N*×*N* were drawn from a generic gamma distribution with mean 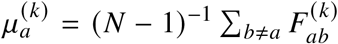 and variance 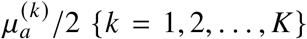. To obtain the unfolded spectra [45], the eigenvalues *E*_*ak*_ are re-scaled as

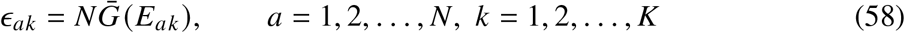

where *G*(*E*) is the empirical level staircase function *G*(*E*) ≡ (*NK*)^−1^ ∑_*ak*_ Θ(*E* − *E*_*ak*_) (*i*.*e*. the cumulative eigenvalue probability function) and 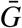 is its smoothed interpolation [52–54]. The nearest level spacings *s*_*ak*_ ≡ *ϵ*_*a*+1,*k*_ − *ϵ*_*a,k*_ are then normalized so that the average level spacing across the entire spectrum is equal to one for all values of *k*:

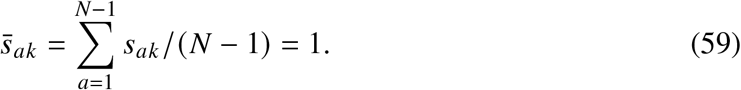

Figure S7**a** shows the level spacing distribution of the unfolded spectrum, 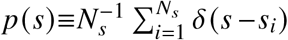, where *N*_*s*_ is the number of independent spacings (for *K* matrices, *N*_*s*_ = *K*(*N* − 1)). The level statistics of the full matrix, Figure S7**a**, is significantly different than the level statistics obtained after all the links of distance larger than 170 km have been omitted (Figure S7**b**). Clearly, the empirical distribution *p*(*s*) interpolates between the Wigner-Dyson distribution of the GOE ensemble (more precisely, the Wigner surmise [45]) that exhibits linear level repulsion typical of extended correlated states:

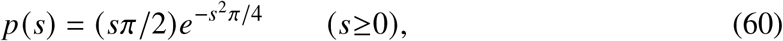

and the Poisson distribution describing independent localized states:

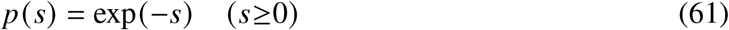

In both equations (60) and (61), the average level spacing is normalized to unity: 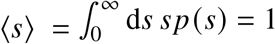.

**Figure S7.**
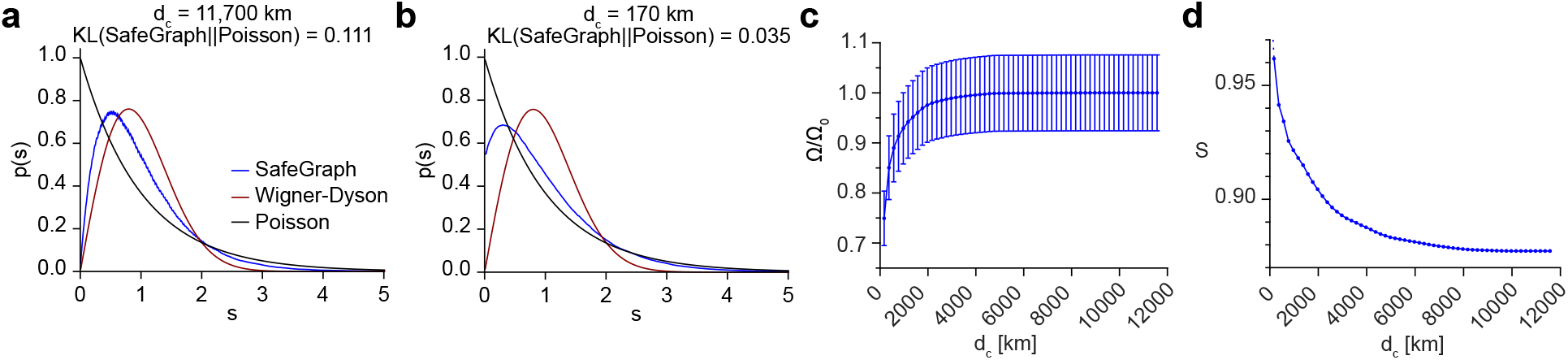
The unfolded level spacing *s*_*i*_ = ε_*i*+1_ − ε_*i*_. **(a)** The level spacing distribution of the full matrix, *i*.*e*. without removing any links. **(b)** The level spacing of the truncated matrix *d*_*c*_ ≤ 170 km. Here 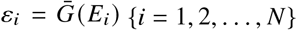, where 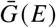 is the smoothed energy staircase function. The GOE level spacing (equation (60)) is plotted in red and the Poisson (equation (61)) in black. **(c)** The relative epidemic size Ω/Ω_0_ as a function of *d*_*c*_. The estimated relative error is ΔΩ/Ω_0_ = 7%. Ω_0_ denotes the unrestricted epidemic size. **(d)** The entropy *S*(SafeGraph) as a function of the cutoff distance *d*_*c*_ in the range 170-12000 km. The maximal value of *d*_*c*_ corresponds to a full flux matrix.

#### 6.2 KL divergence and the level spacing entropy

To quantify the difference between Figure S7**a** and Figure S7**b**, we have calculated the Kullback-Leibler divergence between the empirical spacing distribution, *p*(*s*), and the Poisson distribution

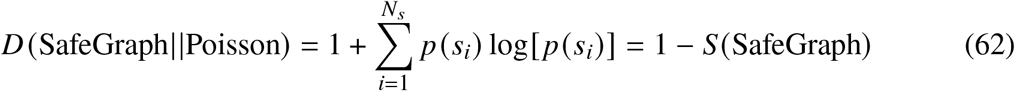

where *D* is the KL divergence and *S* is the entropy of *p*(*s*). The KL divergence of the full matrix is then *D* ≃ 0.11. The KL divergence of the truncated matrix, that shows significantly less level repulsion, and is closer to Poisson statistics, is smaller than that of the full matrix by a factor of three: *D* ≃ 0.04. The KL divergence between the GOE and the Poisson distributions is

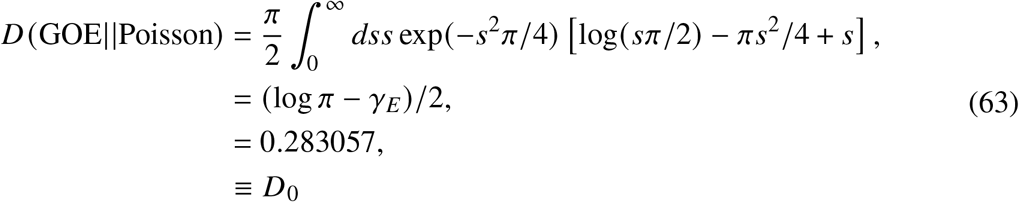

where *γ*_*E*_ = 0.57721 … is the Euler constant. Therefore, in terms relative to *D*_0_ we obtain: *D*/*D*_0_ = {0.39, 0.12} for the full and the truncated matrices respectively. Thus, *D*(SafeGraph||Poisson) is smaller than *D*_0_ and approaches zero as links are being removed. Equivalently, since (i) the entropy of the Poisson distribution is equal to its average ⟨*s*⟩ = 1 and (ii) the entropy of Wigner-Dyson distribution is 1 − *D*_0_ ≃ 0.72, the level spacing entropy varies in the range 1 − *D*_0_ *< S*(SafeGraph)≤1 and it approaches unity as links are removed and the states get localized.

The entropy as a function of the cutoff distance *d*_*c*_ is shown in Figure S7**d**. The distribution *p*(*s*) is found by (i) removing all links of distance larger than *d*_*c*_ (ii) unfolding the spectra of the ensemble of *K* truncated matrices and (iii) repeating the procedure for each value of *d*_*c*_. The corresponding size of the epidemic Ω(∞) and its error ΔΩ are estimated by solving equations (29) and (30) for each one of the truncated matrices and then calculating the mean and standard deviation over *K* samples. In the numerical computation we assumed a recovery rate *γ* = 0.125 d^−1^ and initial fraction of infections *ϵ* = 10^−4^ uniform for all the communities. The epidemic size Ω/Ω_0_ as a function of *d*_*c*_, relative to the value of the unrestricted epidemic size Ω_0_ (*i*.*e*. for the case of a full untruncated matrix), is shown in Figure S7**c**. Thus, a cutoff distance of *d*_*c*_ ≃ 500 km leads to a 10% reduction in the epidemic size. Similarly, *d*_*c*_ ≃ 200 km reduces the epidemic size by 25%. These results are in accordance with the time evolution of the epidemic as is presented in Figure 3**b** of the main text.

#### 6.3 Connectedness and epidemic size

The crossover from Wigner-Dyson to Poisson distribution as links between communities are being successively removed, suggests a policy of isolation which can lead to an effective reduction of the epidemic size. Edges connecting vertices *a* and *b* can be removed in several ways. For example, (i) according to the geographical distance *d*(*a, b*) as has already been done in section 6.2, (ii) according to the “nominal” distance |*a* − *b*|, or (iii) by using the edge-betweenness centrality index, as proposed by Girvan and Newman [49]. Note, that in each one of these cases, one can as well consider a moderated mitigation policy: instead of eliminating links completely, one can impose constraints on the flux of people that are allowed to commute via central pre-determined links.

**Figure S8.**
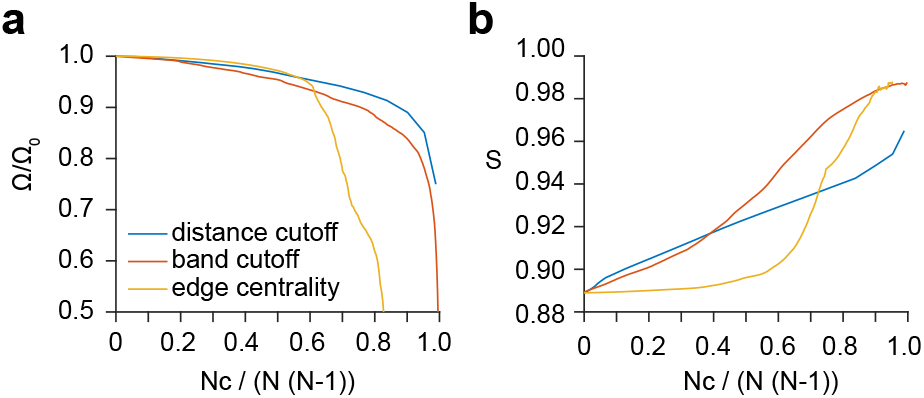
**(a)** The epidemic size Ω/Ω_0_ versus the relative number of cuts *N*_*c*_/*N*/(*N* − 1) for the 3 mitigation methods: distance cutoff, bandwidth, and edge-betweenness centrality. Ω_0_ is the epidemic size for *N*_*c*_ = 0. **(b)** A comparison of the entropy versus the relative number of cuts for the three mitigation method.

To study mitigation according to the nominal distance |*a* − *b*|, we consider the ensemble of banded matrices *F*_*ab*_ with half-bandwidth 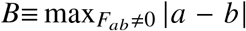, such that the number of non-zero diagonals is 2*B* + 1 irrespective of the underlying physical distance. The increase of Ω/Ω_0_ as a function of *B*, along with a corresponding decrease of the entropy *S*(SafeGraph), is shown in Figure 5**d** of the main text. Comparing to Figure 5**c** of the main text, a reduction of 10% in Ω/Ω_0_ amounts to entropy values of *S* = {0.94, 0.98} for the geographical and nominal cutoffs, respectively. One may argue that mitigation based on “nominal” distance is meaningless, because such a distance changes under the permutations of sites. Indeed, a certain link may either be kept as is, or be omitted, according to an arbitrary re-ordering of communities. However, performing such permutation only reshuffles the matrices of the random ensemble among themselves. Consequently, for *K, N* ≫ 1, the level statistics and the resulting epidemic size are hardly affected.

Centrality indices have been used for detecting the modular structure of social and biological networks [49, 50]. The edge-betweenness centrality of edge (*a, b*) is defined as [51]: 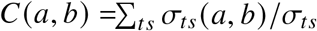, where (*t, s*) stands for “target” and “source” vertices, respectively. Here, *σ*_*ts*_ are shortest paths going *t* ← *s* and *σ*_*ts*_ (*a, b*) are such paths which, in addition, go along edge *a, b*. The entries *σ*_*s*_ are weighted by the flux that they can carry (otherwise, unweighted paths take only binary values *σ*_*ts*_ = 0, 1). The summation is carried over all (*t, s*) pairs that are different from (*a, b*). As observed in [49], edges running between loosely-connected communities should have high values of *C*.

The scheme proposed in [49] for identifying communities in a network is as follows: (i) calculate *C* for all edges of the network (ii) remove the edge with highest *C* (iii) re-calculate *C* for all edges affected by the removal. (iv) Repeat from step (ii) until no edges remain. Here, we are using the same procedure for mitigating the epidemic. There are few practical modifications though:

- Edge-betweenness is calculated for the mean matrix 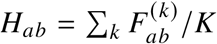. The removal of links is then applied to each matrix separately.
- For a faster computation of step (ii), edges are removed in chunks rather than one-by-one. This is done with the help of the efficient shareware package [MatlabBGL](http://www.mathworks.com/matlabcentral/fileexchange/10922-matlabbgl) that is able to rank all the edges of *H* in a single instance. The initial size of a chunk is 10000 (about 1 % of the total number of links in *H*) and it then decreases adaptively.
- The computation terminates when the resulting KL divergence is sufficiently close to zero. Our modified procedure is clearly sub-optimal. We found, however, that compared to the exhaustive search (i)-(iii), the results almost don’t change because the variations of the KL divergence at the initial stages are relatively small.

The entropy *S*(SafeGraph) and the size of epidemic Ω/Ω_0_ as a function of the number of removed links, *N*_*C*_ (*i*.*e*. number of cuts), are shown in Figure S8. The size of epidemic drops abruptly when the relative number of cuts *C* ≡ *N*_*C*_/*N*/(*N* − 1) exceeds the value *C* = 0.6 (from a relative size of 95 % at *C* = 0.6 to 75 % at *C* = 0.7). This drop is accompanied by the increase of entropy around the same value of *C*. Thus, mitigation based on edge-betweenness performs better than either the distance or the nominal cutoffs, none of which exhibits a similar abrupt crossover. The betweenness index outperforms the other two methods mainly because it is sensitive to the flux that is flowing through links. The “transition” at *c* = 0.6 is expected to become much sharper as *N* ≫ 1 approaches the thermodynamic limit. The evolution of *p*(*s*) as links are gradually being removed is demonstrated in movies S5-S7.

## References

1. Henderson, J. Florence Under Siege: Surviving Plague in an Early Modern City xviii+363 (Yale University Press, New Haven and London, 2019).

2. Kermack, W. O. & McKendrick, A. G. A contribution to the mathematical theory of epidemics. Proceedings of the Royal Society of London. Series A, Containing papers of a mathematical and physical character 115, 700–721 (1927).

3. Hethcote, H. W. The Mathematics of Infectious Diseases. SIAM Rev. 42, 599–653 (2020).

4. Harko, T., Lobo, F. S. & Mak, M. Exact analytical solutions of the Susceptible-Infected-Recovered (SIR) epidemic model and of the SIR model with equal death and birth rates. Applied Mathematics and Computation 236, 184–194 (2014).

5. Wu, J. T., Leung, K. & Leung, G. M. Nowcasting and forecasting the potential domestic and international spread of the 2019-nCoV outbreak originating in Wuhan, China: a modelling study. Lancet 395, 689–697 (2020).

6. Kucharski, A. J., Russell, T. W., Diamond, C., Liu, Y., Edmunds, J., Funk, S. & Eggo, R. M. Early dynamics of transmission and control of COVID-19: a mathematical modelling study. Lancet Infect. Dis. 20, 535–558 (2020).

7. Allard, A., Moore, C., Scarpino, S. V., Althouse, B. M. & Hébert-Dufresne, L. The role of directionality, heterogeneity and correlations in epidemic risk and spread 2020. 2005.11283 [physics.soc-ph].

8. Aleta, A., Martín-Corral, D., Bakker, M. A., Pastore y Piontti, A., Ajelli, M., Litvinova, M., Chinazzi, M., Dean, N. E., Halloran, M. E., Longini, I. M., Pentland, A., Vespignani, A., Moreno, Y. & Moro, E. Quantifying the importance and location of SARS-CoV-2 transmission events in large metropolitan areas. medRxiv (2020).

9. Hébert-Dufresne, L., Althouse, B. M., Scarpino, S. V. & Allard, A. Beyond 0: hetero-geneity in secondary infections and probabilistic epidemic forecasting. Journal of the Royal Society Interface 17, 20200393 (2020).

10. Britton, T., Ball, F. & Trapman, P. A mathematical model reveals the influence of population heterogeneity on herd immunity to SARS-CoV-2. Science 369, 846–849 (2020).

11. Neipel, J., Bauermann, J., Bo, S., Harmon, T. & Jülicher, F. Power-law population heterogeneity governs epidemic waves. PloS one 15, e0239678 (2020).

12. Sun, K., Wang, W., Gao, L., Wang, Y., Luo, K., Ren, L., Zhan, Z., Chen, X., Zhao, S., Huang, Y., Sun, Q., Liu, Z., Litvinova, M., Vespignani, A., Ajelli, M., Viboud, C. & Yu, H. Transmission heterogeneities, kinetics, and controllability of SARS-CoV-2. Science 371 (2021).

13. Kawagoe, K., Rychnovsky, M., Chang, S. Y., Huber, G., Li, L. M., Miller, J., Pnini, R., Veytsman, B. & Yllanes, D. Epidemic dynamics in inhomogeneous populations and the role of superspreaders. Phys. Rev. Research 3, 033283 (Sept. 2021).

14. Pozderac, C. & Skinner, B. Superspreading of SARS-CoV-2 in the USA. PloS one 16, 1–10 (Mar. 2021).

15. Huber, G., Kamb, M., Kawagoe, K., Li, L. M., Veytsman, B., Yllanes, D. & Zigmond, D. A minimal model for household effects in epidemics. Physical Biology 17, 065010 (2020).

16. Aleta, A., Martín-Corral, D., Pastore y Piontti, A., Ajelli, M., Litvinova, M., Chinazzi, M., Dean, N. E., Halloran, M. E., Longini Jr., I. M., Merler, S., Pentland, A., Vespignani, A., Moro, E. & Moreno, Y. Modelling the impact of testing, contact tracing and household quarantine on second waves of COVID-19. Nat. Hum. Behav. 4, 964–971 (2020).

17. Murray, J. Mathematical biology II. Spatial models and biological applications. Springer-Verlag, New York (2003).

18. Postnikov, E. B. & Sokolov, I. M. Continuum description of a contact infection spread in a SIR model. Mathematical Biosciences 208, 205–215 (2007).

19. Brockmann, D. & Helbing, D. The hidden geometry of complex, network-driven contagion phenomena. Science 342, 1337–1342 (2013).

20. Chu, A., Huber, G., McGeever, A., Veytsman, B. & Yllanes, D. A random-walk-based epidemiological model. Sci. Rep. 11, 19308 (2021).

21. Te Vrugt, M., Bickmann, J. & Wittkowski, R. Effects of social distancing and isolation on epidemic spreading modeled via dynamical density functional theory. Nature communications 11, 1–11 (2020).

22. Tsori, Y. & Granek, R. Epidemiological model for the inhomogeneous spatial spreading of COVID-19 and other diseases. PloS one 16, e0246056 (2021).

23. Tizzoni, M., Bajardi, P., Poletto, C., Ramasco, J. J., Balcan, D., Gonçalves, B., Perra, N., Colizza, V. & Vespignani, A. Real-time numerical forecast of global epidemic spreading: case study of 2009 A/H1N1pdm. BMC Medicine 10, 1–31 (2012).

24. Linka, K., Peirlinck, M., Sahli Costabal, F. & Kuhl, E. Outbreak dynamics of COVID-19 in Europe and the effect of travel restrictions. Computer Methods in Biomechanics and Biomedical Engineering 23, 710–717 (2020).

25. Ivorra, B., Ngom, D. & Ramos, A. M. Be-CoDiS: A mathematical model to predict the risk of human diseases spread between countries. Validation and application to the 2014-15 Ebola Virus Disease epidemic. arXiv preprint 1410.6153 (2014).

26. Hsu, S. & Zee, A. Global spread of infectious diseases. Journal of Biological Systems 12, 289–300 (2004).

27. Nande, A., Adlam, B., Sheen, J., Levy, M. Z. & Hill, A. L. Dynamics of COVID-19 under social distancing measures are driven by transmission network structure. PLoS Comput. Biol. 17, e1008684 (2021).

28. Ventura, P. C., Aleta, A., Rodrigues, F. A. & Moreno, Y. Modeling the effects of social distancing on the large-scale spreading of diseases. 2105.09697 (2021).

29. Mayberry, J., Nattestad, M. & Tuttle, A. The Structure of an Outbreak on a College Campus. Mathematics Magazine 94, 83–98 (2021).

30. Chang, S., Pierson, E., Koh, P. W., Gerardin, J., Redbird, B., Grusky, D. & Leskovec, J. Mobility network models of COVID-19 explain inequities and inform reopening. Nature 589, 82–87 (2021).

31. Hethcote, H. W. An immunization model for a heterogeneous population. Theoretical Population Biology 14, 338–349 (1978).

32. Post, W., DeAngelis, D. & Travis, C. Endemic disease in environments with spatially heterogeneous host populations. Mathematical Biosciences 63, 289–302 (1983).

33. May, R. M. & Anderson, R. M. Spatial heterogeneity and the design of immunization programs. Mathematical Biosciences 72, 83–111 (1984).

34. Hethcote, H. W. & Van Ark, J. W. Epidemiological models for heterogeneous populations: proportionate mixing, parameter estimation, and immunization programs. Mathematical Biosciences 84, 85–118 (1987).

35. Lloyd, A. L. & May, R. M. Spatial heterogeneity in epidemic models. Journal of Theoretical Biology 179, 1–11 (1996).

36. SafeGraph. SafeGraph Social Distancing Metrics https://docs.safegraph.com/docs/social-distancing-metrics, year = 2021. 2021.

37. Dong, E., Du, H. & Gardner, L. An interactive web-based dashboard to track COVID-19 in real time. The Lancet infectious diseases 20, 533–534 (2020).

38. Naether, U., Postnikov, E. & Sokolov, I. Infection fronts in contact disease spread. The European Physical Journal B 65, 353–359 (2008).

39. Dee, G. & Langer, J. Propagating pattern selection. Physical Review Letters 50, 383 (1983).

40. Ben-Jacob, E., Brand, H., Dee, G., Kramer, L. & Langer, J. Pattern propagation in nonlinear dissipative systems. Physica D: Nonlinear Phenomena 14, 348–364 (1985).

41. Bramson, M. Convergence of solutions of the Kolmogorov equation to travelling waves (American Mathematical Soc., 1983).

42. Paquette, G., Chen, L.-Y., Goldenfeld, N. & Oono, Y. Structural stability and renormalization group for propagating fronts. Physical review letters 72, 76 (1994).

43. Ebert, U. & van Saarloos, W. Front propagation into unstable states: universal algebraic convergence towards uniformly translating pulled fronts. Physica D: Nonlinear Phenomena 146, 1–99 (2000).

44. Birzu, G., Hallatschek, O. & Korolev, K. S. Fluctuations uncover a distinct class of traveling waves. Proceedings of the National Academy of Sciences 115, E3645–E3654 (2018).

45. Mehta, M. Random Matrices 3rd. Sec. 1.5 (Elsevier, 2004).

46. Akemann, G., Baik, J. & Di Francesco, P. The Oxford handbook of random matrix theory (Oxford University Press, 2011).

47. Livan, G., Novaes, M. & Vivo, P. Introduction to Random Matrices: Theory and Practice (Springer, 2020).

48. Rosenzweig, N. & Porter, C. Repulsion of Energy Levels in Complex Atomic Spectra. Phys. Rev. 120, 1698–1714 (1960).

49. Girvan, M. & Newman, M. E. Community structure in social and biological networks. Proceedings of the National Academy of Sciences 99, 7821–7826 (2002).

50. Luo, F., Zhong, J., Yang, Y., Scheuermann, R. H. & Zhou, J. Application of random matrix theory to biological networks. Physics Letters A 357, 420–423 (2006).

51. DeMeo, P., Ferrara, E., Fiumara, G. & Ricciardello, A. A novel measure of edge centrality in social networks. Jour. Knowledge-Based Systems 30, 136–150 (2012).

52. Brody, T. A., Flores, J., French, J. B., Mello, P., Pandey, A. & Wong, S. S. Random-matrix physics: spectrum and strength fluctuations. Reviews of Modern Physics 53. p. 391, 385– 479 (1981).

53. Guhr, T., Müeller-Groeling, A. & Weidenmüller, H. A. Random matrix theories in quantum physics: common concepts. Phys. Rep. 299. p. 228, 189–425 (1998).

54. Hakke, F. Quantum signatures of Chaos 3rd. Sec. 4.5 (Springer, 2010).

55. Sen, P., Yamana, T. K., Kandula, S., Galanti, M. & Shaman, J. Burden and characteristics of COVID-19 in the United States during 2020. Nature 598, 338–341 (2021).

## Supplementary References

45. Mehta, M. Random Matrices 3rd. Sec. 1.5 (Elsevier, 2004).

54. Hakke, F. Quantum signatures of Chaos 3rd. Sec. 4.5 (Springer, 2010).

